# Specific heterozygous frameshift variants in *hnRNPA2B1* cause early-onset oculopharyngeal muscular dystrophy

**DOI:** 10.1101/2021.04.08.21254942

**Authors:** Hong Joo Kim, Payam Mohassel, Sandra Donkervoort, Lin Guo, Kevin O’Donovan, Maura Coughlin, Xaviere Lornage, Nicola Foulds, Simon R. Hammans, A. Reghan Foley, Charlotte M. Fare, Alice F. Ford, Masashi Ogasawara, Aki Sato, Aritoshi Iida, Pinki Munot, Gautam Ambegaonkar, Rahul Phadke, Dominic G O’Donovan, Rebecca Buchert, Mona Grimmel, Ana Töpf, Irina T. Zaharieva, Lauren Brady, Ying Hu, Thomas E. Lloyd, Andrea Klein, Maja Steinlin, Alice Kuster, Sandra Mercier, Pascale Marcorelles, Yann Péréon, Emmanuelle Fleurence, Adnan Manzur, Sarah Ennis, Rosanna Upstill-Goddard, Luca Bello, Cinzia Bertolin, Elena Pegoraro, Leonardo Salviati, Courtney E. French, Andriy Shatillo, F Lucy Raymond, Tobias Haack, Susana Quijano-Roy, Johann Böhm, Isabelle Nelson, Tanya Stojkovic, Teresinha Evangelista, Volker Straub, Norma B. Romero, Jocelyn Laporte, Francesco Muntoni, Ichizo Nishino, Mark A. Tarnopolsky, James Shorter, J. Paul Taylor, Carsten G. Bönnemann

**Affiliations:** Department of Cell and Molecular Biology, St. Jude Children’s Research Hospital, Memphis, TN, USA; National Institute of Neurological Disorders and Stroke, National Institutes of Health, Bethesda, MD, USA; Department of Biochemistry & Biophysics, Perelman School of Medicine at the University of Pennsylvania, Philadelphia, PA, USA; Department of Biochemistry and Molecular Biology, Thomas Jefferson University, Philadelphia, PA 19107, USA.; Département Médecine Translationnelle et Neurogénétique, Institut de Génétique et de Biologie Moléculaire et Cellulaire, Institut National de la Santé et de la Recherche Médicale U1258, Centre National de la Recherche Scientifique UMR7104, Université de Strasbourg, Illkirch, France; Wessex Clinical Genetics Services, Princess Anne Hospital, Academic Unit of Human Development and Health, Faculty of Medicine, University of Southampton, Southampton, England; Wessex Neurological Centre, University Hospital Southampton, Southampton, UK; Department of Neuromuscular Research, National Institute of Neuroscience, National Center of Neurology and Psychiatry (NCNP), 4-1-1 Ogawahigashi, Kodaira, Tokyo 187-8502, Japan; Medical Genome Center, NCNP, Kodaira, Tokyo, Japan; Department of Neurology, Niigata City General Hospital, Niigata, Japan; The Dubowitz Neuromuscular Centre, NIHR Great Ormond Street Hospital Biomedical Research Centre, Great Ormond Street Institute of Child Health, University College London, & Great Ormond Street Hospital Trust, London, UK; Department of Paediatric Neurology, Cambridge University Hospital NHS Trust, Addenbrookes Hospital, Cambridge CB2 0QQ, UK; Division of Neuropathology, University College London Hospitals NHS Foundation Trust National Hospital for Neurology and Neurosurgery London, UK and Division of Neuropathology, UCL Institute of Neurology, Dubowitz Neuromuscular Centre, London, UK; Department of Histopathology Box 235, Level 5 John Bonnett Clinical Laboratories Addenbrooke’s Hospital, Cambridge, UK; Institute of Medical Genetics and Applied Genomics, University of Tübingen, Tübingen, Germany; John Walton Muscular Dystrophy Research Centre, Newcastle University and Newcastle Hospitals NHS Foundation Trust, Newcastle upon Tyne, UK; Division of Neuromuscular & Neurometabolic Disorders, Department of Pediatrics, McMaster University, Hamilton Health Sciences Centre, Hamilton, ON, Canada; Department of Neurology, Johns Hopkins University School of Medicine, Baltimore, MD, USA; Division of Neuropaediatrics, Development and Rehabilitation, Children’s University Hospital, Inselspital, Bern, Switzerland; Pediatric Neurology, University Children’s Hospital Basel, University of Basel, Basel, Switzerland; Department of Neurometabolism, University Hospital of Nantes, Nantes, France; CHU de Nantes, Service de Génétique Médicale, Centre de Référence des Maladies Neuromusculaires, Hôtel-Dieu, Nantes, France; L’institut du thorax, INSERM, CNRS, UNIV Nantes, Nantes, France; Service d’anatomopathologie, CHU de Brest, Hôpital Morvan, Brest Cedex, France. EA 4685 LIEN, Université de Bretagne Occidentale, Brest Cedex, France; CHU de Nantes, Centre de Référence des Maladies Neuromusculaires, Hôtel-Dieu, Nantes, France; Etablissement de Santé pour Enfants et Adolescents de la région Nantaise, Nantes, France; Human Genetics and Genomic Medicine, Faculty of Medicine, University of Southampton, Southampton, UK; Department of Neurosciences, DNS, University of Padova, Padova, Italy; Clinical Genetics Unit, Department of Women and Children’s Health, University of Padova, Padova, Italy and IRP Città della Speranza, Padova, Italy; Department of Paediatrics, University of Cambridge, Cambridge, UK; Institute of Neurology, Psychiatry and Narcology of NAMS of Ukraine, Kharkiv, Ukraine; Cambridge Institute of Medical Research, University of Cambridge, Cambridge, UK; Neuromuscular Unit, Pediatric Neurology and ICU Department, Raymond Poincaré Hospital (UVSQ), AP-HP Université Paris-Saclay, Garches, France; Sorbonne Université, INSERM, Centre of Research in Myology, UMRS974, Paris, France; Centre de référence des Maladies Neuromusculaires Nord/Est/Ile de France, Institut de Myologie, Sorbonne Université, Hôpital Pitié-Salpêtrière, Paris, France; Unité de Morphologie Neuromusculaire, Institut de Myologie, Sorbonne Université, Hôpital Pitié-Salpêtrière, Paris, France; Howard Hughes Medical Institute, Chevy Chase, MD, USA

## Abstract

RNA-binding proteins (RBPs) are essential for post-transcriptional regulation and processing of RNAs. Pathogenic missense variants in RBPs underlie a spectrum of disease phenotypes, including amyotrophic lateral sclerosis, frontotemporal dementia, inclusion body myopathy, distal myopathy, and Paget’s disease of the bone. Here, we present ten independent families with a severe, progressive, early-onset muscular dystrophy, reminiscent of oculopharyngeal muscular dystrophy (OPMD), caused by heterozygous frameshift variants in the prion-like domain of *hnRNPA2B1*. We found that in contrast with the previously reported missense variants, the frameshift *hnRNPA2B1* variants do not promote, but rather decelerate the fibrillization of the protein. Importantly, the frameshift variants harbor altered nuclear-localization sequences and exhibit reduced affinity for the nuclear-import receptor, Karyopherin-β2, which promotes their cytoplasmic accumulation in cells and in animal models that recapitulate the human pathology. Thus, we expand the phenotypes associated with *hnRNPA2B1* to include a severe, early-onset disease reminiscent of OPMD, caused by a distinct class of frameshift variants that alter its nucleocytoplasmic transport dynamics.

## Main

RNA-binding proteins (RBPs) play central roles in post-transcriptional regulation of RNAs, including RNA splicing, polyadenylation, stabilization, localization and translation^1^. Under certain physiologic conditions, RBPs form membraneless organelles such as nucleoli and stress granules via liquid-liquid phase separation (LLPS)^2^. Most RBPs contain low complexity domains (LCDs) that similar to yeast prion domains are enriched in glycine and uncharged polar amino acids and can be more specifically referred to as prion-like domains (PrLDs). These PrLDs contribute to LLPS by participating in weak multivalent interactions, which may be important for optimal RBP function^3^.

Pathogenic missense variants in RBPs such as TDP-43^4,5^, hnRNPA1^6^, hnRNPA2B1^6^, FUS^7,8^ and TIA1^9,10^ cause a spectrum of diseases with pleomorphic phenotypic manifestations including amyotrophic lateral sclerosis (ALS)/motor neuron disease, frontotemporal dementia (FTD), inclusion body myopathy (IBM), distal myopathy, and Paget’s disease of the bone (PDB). Patients may also present with complex phenotypes impacting muscle, brain and bone, a syndrome that has been termed multisystem proteinopathy (MSP)^11^. Remarkably, most pathogenic variants in RBPs are located in PrLDs^4–10^. Studies using purified proteins and cellular expression models have demonstrated that PrLD variants can alter LLPS and ultimately result in aggregation and fibrillization of the mutant protein^6,9^.

We previously reported a rare MSP phenotype manifesting as PDB, IBM, ALS, and FTD caused by a *hnRNPA2B1* [NM_002137, p.(D290V)] missense variant^6^. This variant, which is within the PrLD of the protein, promotes assembly into self-seeding fibrils by introducing a potent steric-zipper motif, thereby dysregulating polymerization, and ultimately driving the formation of cytoplasmic inclusions^6,12^. In addition, a heterozygous *hnRNPA2B1* [NM_002137, p.(P298L)] missense variant, also within the PrLD and predicted to promote aggregate formation, has been reported to cause PDB without additional phenotypic features^13^. The distinct molecular and mechanistic events underlying the pleomorphic phenotypes of *hnRNPA2B1-*related disease and how they relate to relevant cell types remain unclear.

Oculopharyngeal muscular dystrophy (OPMD) is another disease related to variants in an RBP. Trinucleotide expansions in *PABPN1* underlie the majority of OPMD cases, a late-onset (typically 5^th^ decade) disease characterized by ptosis, dysphagia and, in rare cases, progressive limb weakness and in later stages, vertical ophthalmoplegia^14^. OPMD is distinct from the pleomorphic syndrome MSP, and, unlike other RBP-associated diseases, has a homogeneous and recognizable phenotypic spectrum that predominantly manifests in select skeletal muscles.

Here, we present and characterize ten independent families with a severe, progressive, early-onset OPMD-like phenotype that is distinct from MSP and is caused by a novel class of heterozygous frameshift variants in the PrLD of *hnRNPA2B1*. In contrast to the missense variants that cause MSP or PDB, these frameshift variants do not promote but rather decelerate protein fibrillization. The frameshift variants alter the C-terminal portion of the nuclear-localization sequence and promote cytoplasmic accumulation of the protein by impairing its interaction with its nuclear-import receptor, Karyopherin-β2 (Kapβ2). Thus, this study expands the clinical phenotypes associated with *hnRNPA2B1* to include a severe, early-onset disease reminiscent of OPMD caused by a distinct class of frameshift variants with specific consequences on hnRNPA2B1 nucleocytoplasmic transport dynamics.

## Results

### *hnRNPA2B1* frameshift variants manifest with a distinct, early-onset progressive myopathy

We identified eleven patients from ten independent families with progressive early-onset myopathy with ophthalmoplegia, ptosis and respiratory insufficiency of variable degrees (Table 1, Fig. 1a). Six patients presented with first recognition of symptoms before two years of age, ranging from respiratory insufficiency requiring tracheostomy at birth, to delayed motor milestones or isolated ptosis and ophthalmoplegia. All patients had axial weakness and progressive proximal and distal weakness, which was more pronounced in the lower extremities compared to the upper extremities (data not shown). Respiratory involvement was variable, ranging from a severely decreased forced vital capacity (FVC) (21% predicted) to normal. Ptosis and ophthalmoplegia were uniformly present in all patients and were noted as early as six months of age. Cognition was normal, and patients did not have a history of seizures, cardiac involvement, or bone abnormalities. Serum creatine kinase levels were elevated in all patients, ranging from 3 to 35 times the upper limit of normal (Table 1). Detailed clinical and genetic information for each patient can be found in Supplementary Table 1 and Supplementary Information.

**Table 1.**
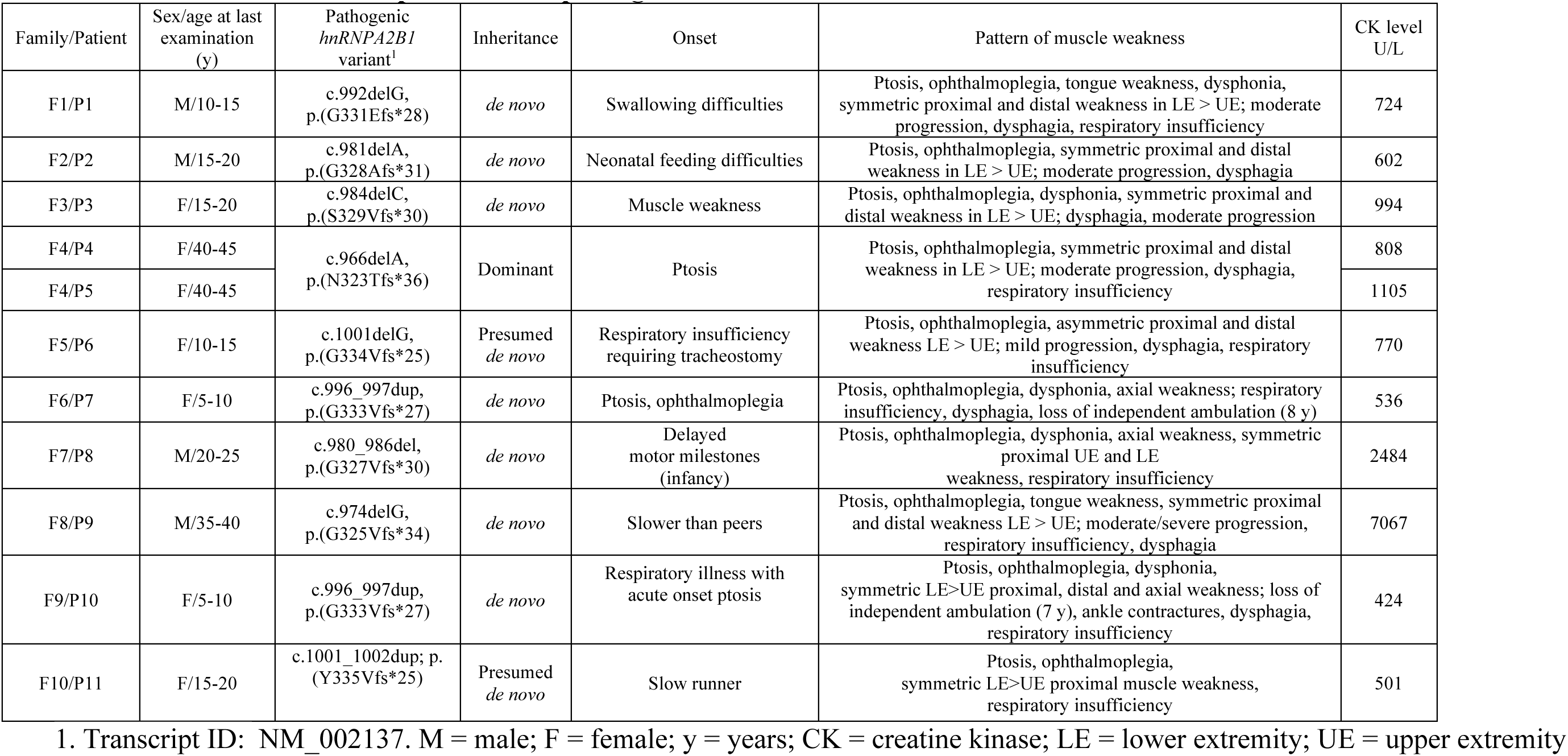
Clinical features of patients with pathogenic variants in *hnRNPA2B1*.

| Family/Patient | Sex/age at last examination (y) | Pathogenic *hnRNPA2B1* variant<sup>1</sup> | Inheritance | Onset | Pattern of muscle weakness | CK level U/L |
| --- | --- | --- | --- | --- | --- | --- |
| F1/P1 | M/10-15 | c.992delG, p.(G331Efs*28) | *de novo* | Swallowing difficulties | Ptosis, ophthalmoplegia, tongue weakness, dysphonia, symmetric proximal and distal weakness in LE > UE; moderate progression, dysphagia, respiratory insufficiency | 724 |
| F2/P2 | M/15-20 | c.981delA, p.(G328Afs*31) | *de novo* | Neonatal feeding difficulties | Ptosis, ophthalmoplegia, symmetric proximal and distal weakness in LE > UE; moderate progression, dysphagia | 602 |
| F3/P3 | F/15-20 | c.984delC, p.(S329Vfs*30) | *de novo* | Muscle weakness | Ptosis, ophthalmoplegia, dysphonia, symmetric proximal and distal weakness in LE > UE; dysphagia, moderate progression | 994 |
| F4/P4 | F/40-45 | c.966delA, p.(N323Tfs*36) | Dominant | Ptosis | Ptosis, ophthalmoplegia, symmetric proximal and distal weakness in LE > UE; moderate progression, dysphagia, respiratory insufficiency | 808 |
| F4/P5 | F/40-45 |  |  |  |  | 1105 |
| F5/P6 | F/10-15 | c.1001delG, p.(G334Vfs*25) | Presumed *de novo* | Respiratory insufficiency requiring tracheostomy | Ptosis, ophthalmoplegia, asymmetric proximal and distal weakness LE > UE; mild progression, dysphagia, respiratory insufficiency | 770 |
| F6/P7 | F/5-10 | c.996_997dup, p.(G333Vfs*27) | *de novo* | Ptosis, ophthalmoplegia | Ptosis, ophthalmoplegia, dysphonia, axial weakness; respiratory insufficiency, dysphagia, loss of independent ambulation (8 y) | 536 |
| F7/P8 | M/20-25 | c.980_986del, p.(G327Vfs*30) | *de novo* | Delayed motor milestones (infancy) | Ptosis, ophthalmoplegia, dysphonia, axial weakness, symmetric proximal UE and LE weakness, respiratory insufficiency | 2484 |
| F8/P9 | M/35-40 | c.974delG, p.(G325Vfs*34) | *de novo* | Slower than peers | Ptosis, ophthalmoplegia, tongue weakness, symmetric proximal and distal weakness LE > UE; moderate/severe progression, respiratory insufficiency, dysphagia | 7067 |
| F9/P10 | F/5-10 | c.996_997dup, p.(G333Vfs*27) | *de novo* | Respiratory illness with acute onset ptosis | Ptosis, ophthalmoplegia, dysphonia, symmetric LE>UE proximal, distal and axial weakness; loss of independent ambulation (7 y), ankle contractures, dysphagia, respiratory insufficiency | 424 |
| F10/P11 | F/15-20 | c.1001_1002dup; p.(Y335Vfs*25) | Presumed *de novo* | Slow runner | Ptosis, ophthalmoplegia, symmetric LE>UE proximal muscle weakness, respiratory insufficiency | 501 |

1. Transcript ID: NM\_002137. M = male; F = female; y = years; CK = creatine kinase; LE = lower extremity; UE = upper extremity

**Fig. 1:**
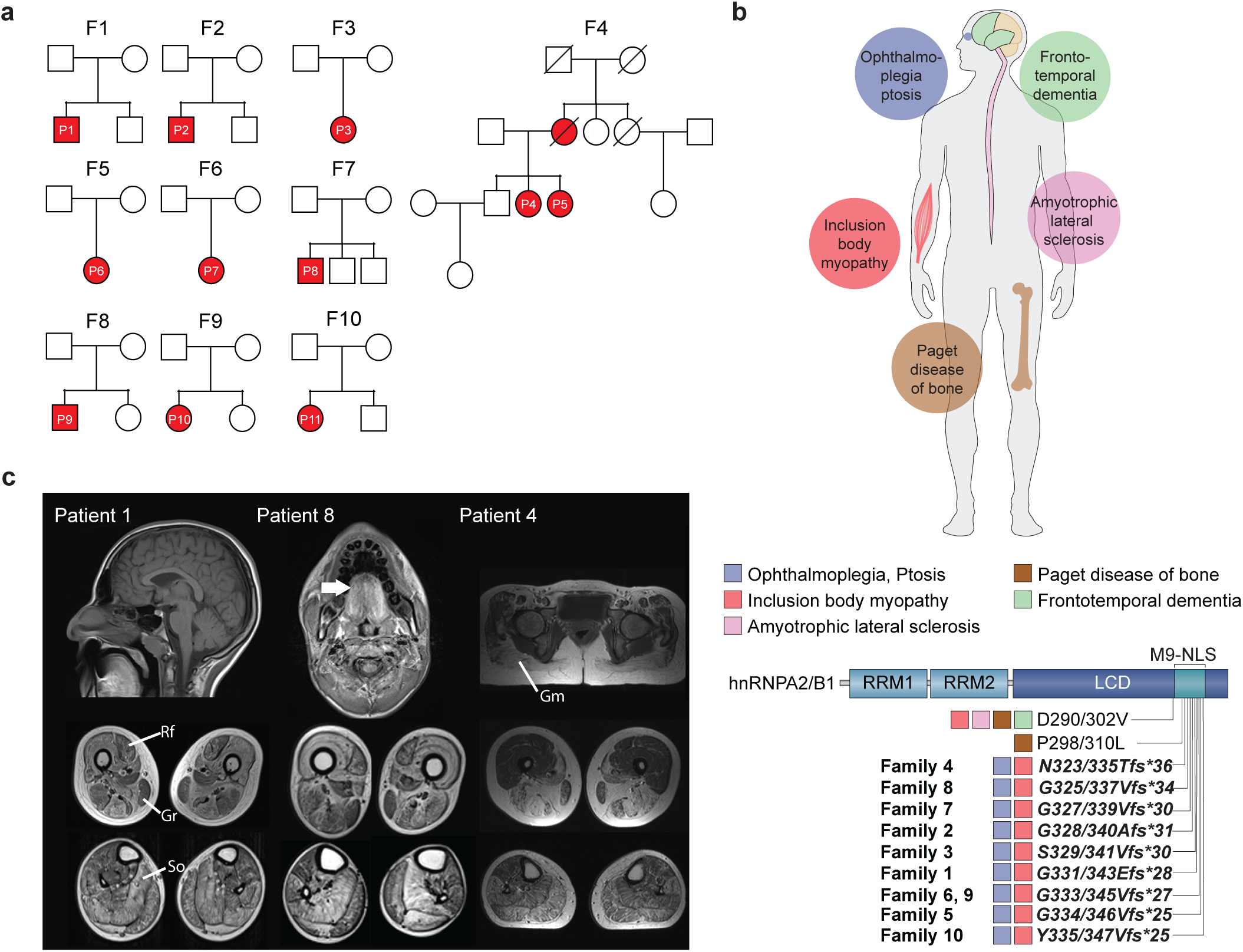
Clinical characteristics and genetic findings in ten independent families with *hnRNPA2B1* variants. **a,** Pedigrees of the ten families. F=family, P=patient **b,** hnRNPA2B1 domain structure with conserved regions. Variants identified from previous studies and in this study (in bold) are indicated in both hnRNPA2 and hnRNPB1 isoforms with their associated phenotypic features. RRM: RNA-recognition motif; LCD: low complexity domain; M9-NLS: M9-nuclear localization signal. **c,** T1 MRI images of the head and lower extremities. Head and neck MRI highlights T1 hyperintensity in the tongue (arrows indicate “bright tongue sign”). In the hips, gluteus maximus (Gm) is selectively affected. In the lower extremities, there is selective involvement of the posterior and medial thigh compartment with relative sparing of the rectus femoris (Rf) and gracilis (Gr) muscle. Lower leg images show relative sparing of the tibialis anterior and selective involvement of the peroneus group (Pr), soleus (So), and lateral gastrocnemius (Lat G).

To elucidate the genetic origin of the muscle disease, we pursued whole exome sequencing and identified nine (one recurrent) heterozygous frameshift variants in *hnRNPA2B1* (MIM:600124), which are rare and not present in the Genome Aggregation Database (gnomAD)^15^ (Fig. 1b and Supplementary Table 1). In family 4, the disease was inherited in a dominant fashion and the variant segregated with disease. The disease occurred sporadically in the remaining nine families (Fig. 1a). The *hnRNPA2B1* variants were confirmed to be *de novo* (i.e., absent in unaffected parents) in seven families for which parental samples were available for testing.

### Muscle imaging reveals myopathic changes with discrete foci of increased T1 signal

Muscle MRI images of lower extremities showed muscle atrophy and a heterogenous pattern of T1 signal hyperintensity, suggestive of architectural changes in the muscle composition (Fig. 1c). Presence of patchy foci of T1 signal hyperintensity, suggestive of focal fatty replacement of muscle, was similar to what has been reported for related disorders such as *VCP*-related MSP^16^. In the anterior thigh compartment, the rectus femoris muscle was relatively spared. Medial and posterior thigh compartments were selectively affected, albeit not uniformly. Gracilis muscle was also relatively spared. In the lower legs, peroneus group, soleus, and lateral gastrocnemius muscles were selectively affected; however, the prominence of this pattern was variable among patients. In the head and neck MRI images, T1 hyperintensity in the tongue was notable (Fig. 1c, arrows).

### *hnRNPA2B1* frameshift variants manifest pathologically with rimmed vacuoles and intracytoplasmic aggregates

Muscle biopsy findings of the patients were consistent with a chronic myopathy, characterized by myofiber atrophy, fiber size variability, increased internalized nuclei, and presence of subsarcolemmal and cytoplasmic rimmed vacuoles without inflammatory infiltrates (Fig. 2a,b). Light microscopy revealed that the vacuoles contained proteinaceous material and were likely of lysosomal or autophagic origin (Fig 2c-f). Ultrastructural analysis confirmed the presence of membrane-bound autophagic vacuoles containing myelin-like debris (Fig. 2g,h). In addition, cytoplasmic, perinuclear, and on occasion intranuclear tubulofilamentous inclusions with ∼15-20 nm thickness were identified, likely representing microfibrillar protein aggregates (Fig. 2i,j, arrows and inset). Immunofluorescence staining of muscle tissue highlighted scattered myofibers with hnRNPA2B1-positive inclusions that partially colocalized with P62, ubiquilin 2, TIA1 and TDP-43, and to a lesser degree with ubiquitin (Fig. 2k-o), similar to the end-stage fibrillar accumulation of RBPs in microfibrillar structures observed in related disorders^6^. These inclusions did not stain positive for Z-disk associated proteins that typically aggregate in myofibrillar myopathies (Supplementary Fig. 1) and were unvested within membrane layers, suggesting ongoing autophagic degradation.

**Fig. 2:**
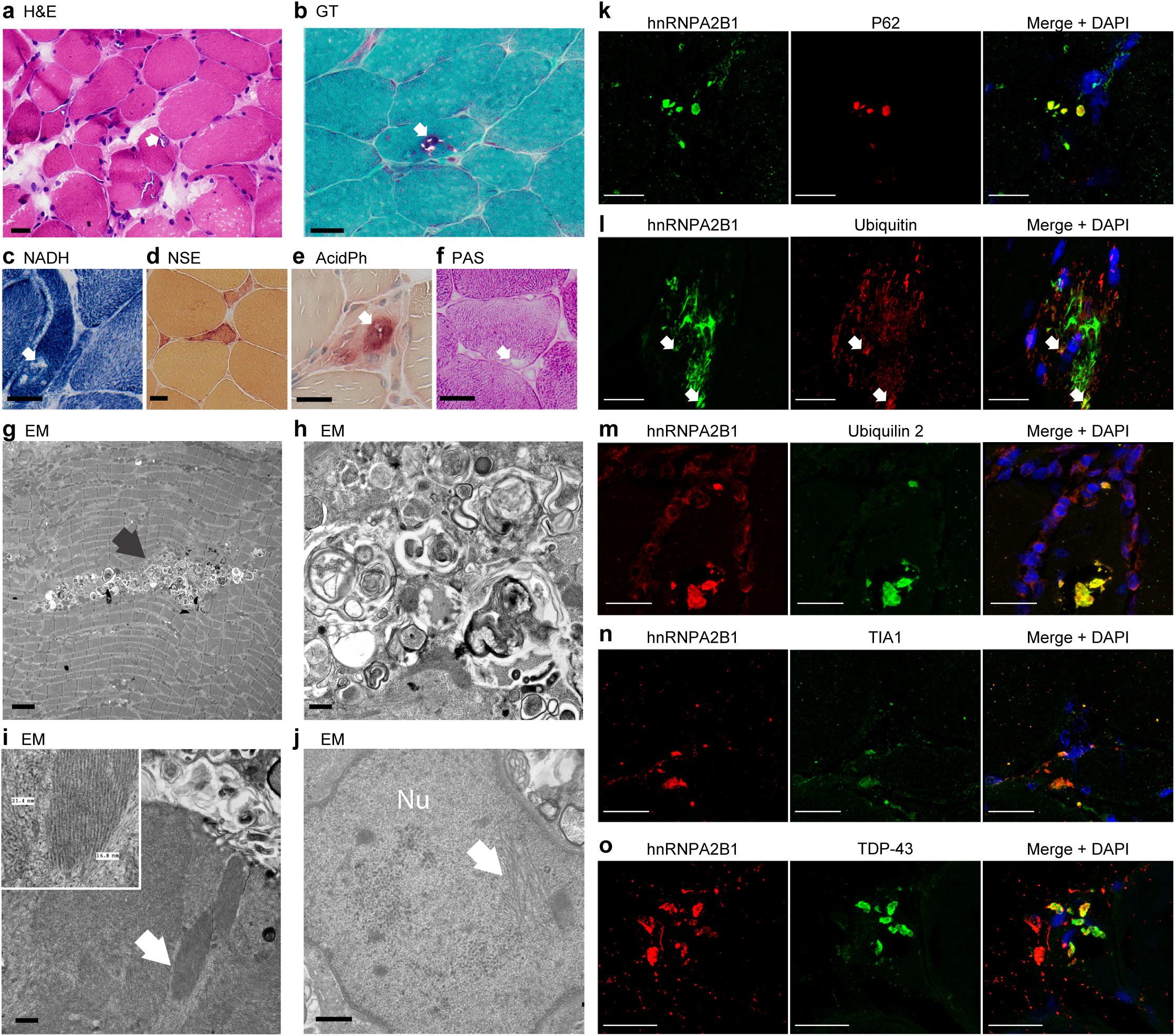
Histological findings of hnRNPA2B1 myopathy. **a**, Hematoxylin and eosin (H&E) image of right rectus femoris muscle biopsy obtained at 8 years of age from patient 1 with c.992delG, p.(G331Efs*28) showing myofiber atrophy, fiber size variability and internalized nuclei with several fibers with rimmed subsarcolemmal vacuoles (arrow). **b,** Modified Gömöri trichrome (GT) staining highlights similar findings and rimmed vacuoles (arrow) (patient 1 muscle biopsy) **c-f,** Vacuole contents (arrow) do not stain positive with NADH, suggesting they are devoid of mitochondria (**c**) and do not show increased non-specific esterase (NSE) activity (**d**). The vacuoles (arrow) do not contain glycoproteins based on periodic acid–Schiff (PAS) stain (**f**) but have increased acid phosphatase (AcidPh) activity (arrow) (**e**), suggesting they are lysosomal or autophagic in origin. NSE stain (**d**) also highlights a few angular atrophic fibers, suggestive of mild acute neurogenic atrophy. **c-f** are from patient 2 muscle biopsy. **g-h,** Electron microscopy (EM) studies showing marked autophagic changes (black arrow) and vacuoles containing membranous myelin-like whorls. **i-j,** Many myofibers contain areas with ∼15-20-nm thick, tubulofilamentous inclusions (**i**, white arrow and inset), which on occasion were also seen within the nuclei (Nu) near the vacuoles (j, white arrow). **g-i** are from patient 2 muscle biopsy, **j** is from patient 8 muscle biopsy **k-o,** Immunofluorescence staining of muscle biopsy of patient 1 with c.992delG, p.(G331Efs*28) showing cytoplasmic hnRNPA2B1-positive inclusions that partially co-localize with P62 (**k**), ubiquilin 2 (**m**), TIA1 (**n**), and TDP-43 (**o**). Co-localization with ubiquitin (**l**) is restricted to very few perinuclear and cytoplasmic puncta (arrows). Scale bars: a-f, 25 µm; g, 2 µm; h-j, 500 nm; k-o, 20 µm.

### Frameshift variants cluster in the *hnRNPA2B1* PrLD and escape RNA quality control degradation

*hnRNPA2B1* is expressed as two alternatively spliced isoforms, hnRNPA2 and hnRNPB1. The shorter hnRNPA2, which lacks exon 2 and its associated 12 amino acids in the N-terminal region, is the main isoform accounting for 90% of the protein in most tissues. All frameshift variants identified in our cohort cluster in the highly conserved PrLD of *hnRNPA2B1* and are predicted to abolish the stop codon, extend the reading frame, and result in expression of the same novel rearranged amino acid sequence at the C-terminal end of both isoforms (Supplementary Fig. 2a,b). RT-PCR analysis in muscle tissue from patients 1 and 2 suggest that these frameshift variants escape RNA quality control degradation (Supplementary Fig. 2c).

### Frameshift variants alter the nucleocytoplasmic ratio of hnRNPA2 and enhance its recruitment to RNA granules

Nucleocytoplasmic shuttling of hnRNPA2B1 is regulated by its 40-amino acid M9 sequence located within the C-terminal LCD (Fig. 3a,b). The M9 sequence serves as both an nuclear localization signal (NLS) and a nuclear export signal (NES) and is recognized by its nuclear transport receptor karyopherin β2 (Kapβ2, also known as transportin 1, TNPO1)^17–19^. M9-NLSs are structurally disordered and have an overall basic character with weakly conserved sequence motifs composed of an N-terminal hydrophobic or basic motif followed by a C-terminal R/H/KX_2-5_PY consensus sequence^20^ (Fig. 3b). Due to the strict conservation of proline-tyrosine (PY) residues and the importance of these residues in the binding of Kapβ2, M9-NLSs have been reclassified as more minimal PY-NLSs, which end on the PY motif^20^.

**Fig. 3:**
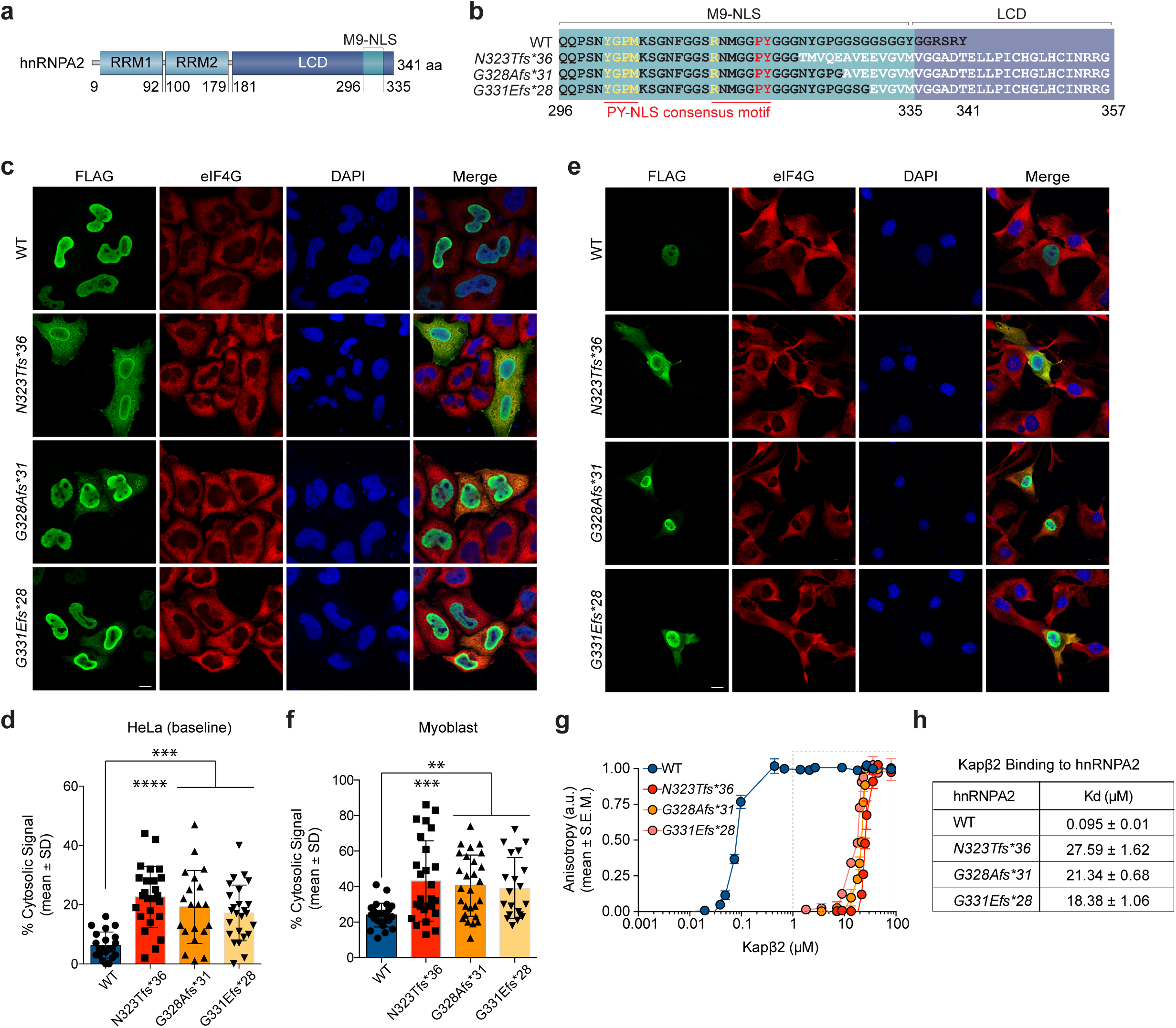
Frameshift mutations impair nucleocytoplasmic trafficking of hnRNPA2 by disrupting interaction between hnRNPA2 and its nuclear transport receptor Kapβ2. **a,** Domain architecture of hnRNPA2. hnRNPA2 contains N-terminal RNA recognition motifs 1 and 2 (RRM1 and RRM2), a C-terminal low complexity domain (LCD) and an M9 nuclear localization signal (M9-NLS) within the LCD. **b,** Amino acid sequences of the relevant domains in WT and each frameshift mutant. The consensus PY-NLS motifs within the M9-NLS are underlined in red. **c,** Intracellular localization of FLAG-tagged hnRNPA2 WT, N323Tfs*36, G328Afs*31, and G331Efs*28 mutants under basal conditions in HeLa cells. eIF4G was used as a cytoplasmic and stress granule marker. Scale bar, 10 μm. **d,** Quantification of hnRNPA2 cytosolic signal intensity in HeLa cells as shown in (c). An interleaved scatter plot with individual data points is shown; error bars represent mean ± s.d. For WT, N323Tfs*36, G328Afs*31, and G331Efs*28 mutants, n=21, 25, 20, and 25 cells, respectively. \*\*\**p* < 0.001, *\*\*\*\*p* < 0.0001 by one-way ANOVA with Dunnett’s multiple comparisons test. **e,** Intracellular localization of FLAG-tagged hnRNPA2 WT, N323Tfs*36, G328Afs*31, and G331Efs*28 mutants in C2C12 myoblasts. eIF4G was used as a cytoplasmic and stress granule marker. Scale bar, 10 μm. **f,** Quantification of hnRNPA2 cytosolic signal intensity in C2C12 myoblasts as shown in (e). An interleaved scatter plot with individual data points is shown; error bars represent mean ± s.d. n=26, 26, 27, and 20 cells for WT, N323Tfs*36, G328Afs*31, and G331Efs*28 mutants, respectively. \*\**p* < 0.01 and \*\*\**p* < 0.001 by two-way ANOVA with Dunnett’s multiple comparisons test. **g,** Fluorescence polarization measurements between TAMRA-labeled M9-NLS of hnRNPA2 WT, N323Tfs*36, G328Afs*31, or G331Efs*28 peptide (100 nM) with increasing concentrations of Kapβ2 WT. Peptide sequences are shown in (b). Values represent means ± s.e.m. (n = 3). **h,** K_d_ values between TAMRA-labeled M9-NLS of hnRNPA2 peptides and Kapβ2 WT. Values were calculated by fitting the curve to the function 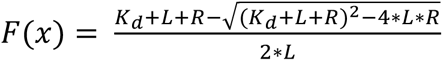, where *K_d_* is the binding affinity, *L* is the concentration of peptide, and *R* is the concentration of Kapβ2. Values represent means ± s.d. from 3 independent experiments.

All nine *hnRNPA2B1* frameshift variants identified in our cohort alter the M9-NLS amino acid sequence by shifting the reading frame by one base pair (Supplementary Fig. 2a,b), suggesting that nucleocytoplasmic trafficking might be impaired by these variants. To examine the effect of these C-terminal frameshift variants on nucleocytoplasmic trafficking of hnRNPA2B1, we expressed FLAG-tagged versions of wild-type (WT) and three different mutant hnRNPA2 proteins (N323Tfs*36, G328Afs*31, and G331Efs*28) in HeLa cells. Whereas hnRNPA2 WT almost exclusively localized to nuclei, N323Tfs*36, G328Afs*31, and G331Efs*28 mutants showed cytoplasmic accumulation at baseline, suggesting impairment of nucleocytoplasmic transport (Fig. 3c,d). The cytoplasmic accumulation of mutant proteins became more apparent when cells were subjected to oxidative stress, which induces assembly of stress granules (Supplementary Fig. 3a,b). We also evaluated the impact of the variants on hnRNPA2 localization in a disease-relevant cell type and found that, similar to HeLa cells, C2C12 myoblast cells (Fig. 3e, f) and differentiated myotubes (Supplementary Fig. 3c) showed increased cytoplasmic localization of mutant hnRNPA2 proteins compared to WT.

### Frameshift variants impair the interaction between hnRNPA2 and its nuclear transport receptor Kapβ2

The conserved consensus motifs of the PY-NLS are located proximal to the region in which the *hnRNPA2B1* reading frame is altered by the frameshift variants (Fig. 3b and Supplementary Fig. 2b), suggesting that the cytoplasmic accumulation of mutant hnRNPA2 might be caused by loss of Kapβ2 interaction and subsequent impaired nuclear import. To test the impact of frameshift variants on Kapβ2 binding, we synthesized WT and three frameshift mutant peptides (Fig. 3b) with a 5′ tetramethylrhodamine (TAMRA) label and used fluorescence polarization assays to quantify their interaction with Kapβ2^21^. The hnRNPA2 WT peptide bound Kapβ2 with a *K_d_* of 95 nM (Fig. 3g,h). All three frameshift mutant peptides showed a significant decrease in Kapβ2 binding (Fig. 3g,h), suggesting that frameshift variants affect the ability of hnRNPA2 to bind Kapβ2 and thus impair its subsequent nuclear import.

### Frameshift variants cause apoptotic cell death in differentiating cells

We observed that upon differentiation, the number of C2C12 cells expressing mutant forms of hnRNPA2 decreased dramatically over time (Fig. 4a,b). Thus, we assessed whether mutant forms of hnRNPA2 caused cell death in differentiating C2C12 cells. Indeed, upon differentiating C2C12 myoblasts into myotubes, increasing numbers of cells expressing GFP or GFP-hnRNPA2 became positive for annexin V and propidium iodide (PI) staining, suggesting that they underwent apoptotic cell death (Fig. 4c,d). Notably, cell death was enhanced in the presence of the frameshift variants, with approximately 60% and 80% of frameshift mutant hnRNPA2-expressing cells becoming positive for annexin V and PI staining one and two days after differentiation, respectively (Fig. 4c,d). Although the overall population of cells expressing hnRNPA2 WT did not show increased cell death compared to GFP-expressing cells at low and medium expression levels (Fig. 4c,d), they showed decreased survival rates at high expression levels of exogenous hnRNPA2 WT (Fig. 4e). Cell toxicity was not enhanced in proportion to expression levels in cells expressing frameshift mutant forms of hnRNPA2, but was most similar to cells expressing high levels of hnRNPA2 WT, suggesting that frameshift-associated cell toxicity could be partially attributable to a gain of wild-type function (Fig. 4e).

**Fig. 4:**
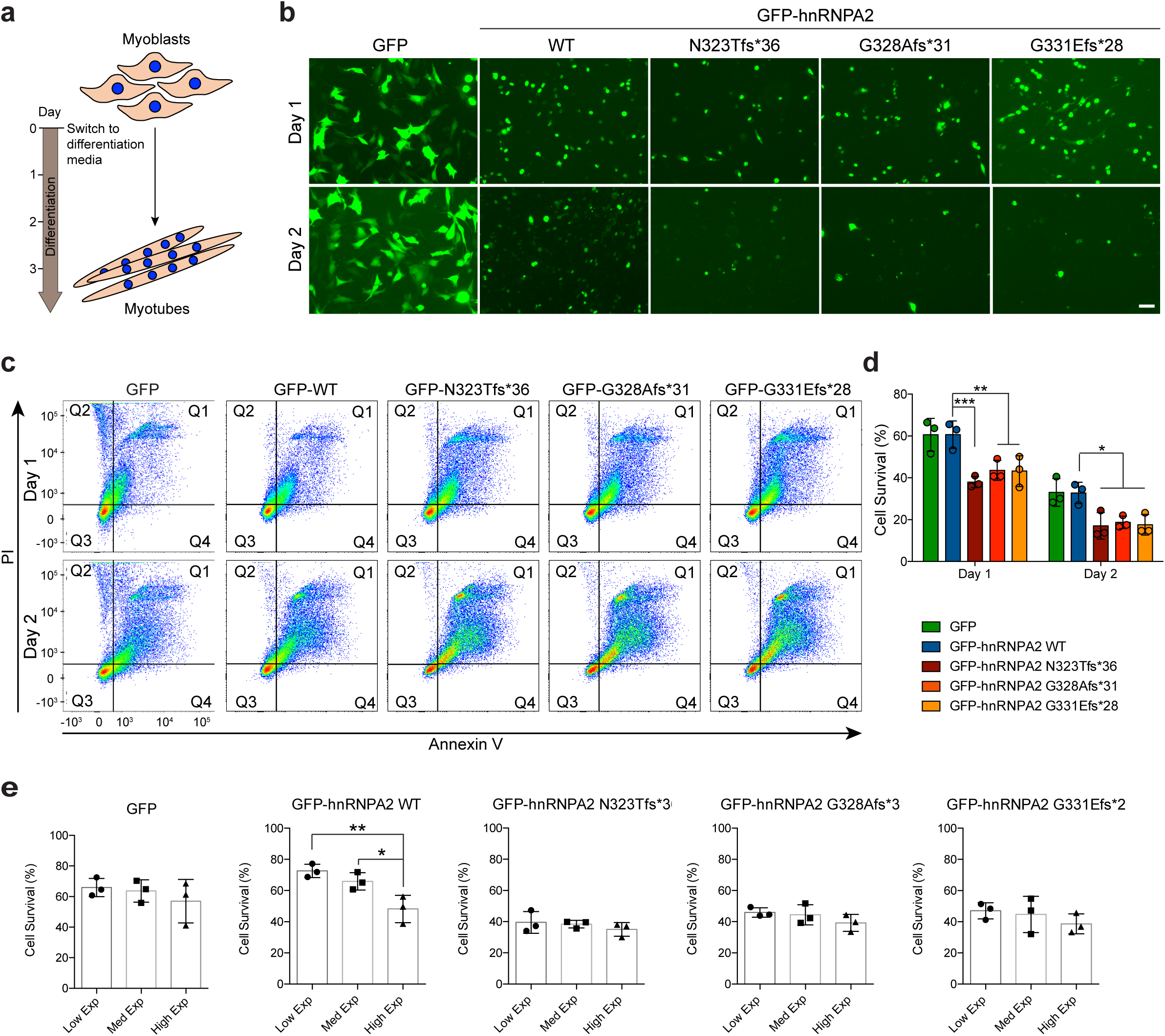
Expression of frameshift variants causes increased cell toxicity during C2C12 differentiation. **a,** Schematic of C2C12 differentiation. Differentiation media consists of DMEM supplemented with 0.5% FBS, 1% penicillin/streptomycin, and 1% L-glutamate. **b,** Representative images of C2C12 cells one (top) or two (bottom) days after change to differentiation media. Scale bar, 20 μm. **c,** Representative scatter plots of GFP-positive C2C12 cells stained with propidium iodide (PI) and annexin V, one (top) or two (bottom) days after change to differentiation media. **d,** Quantification of GFP-positive cells that were negative for both PI and annexin V. Values represent means ± s.e.m. (n = 3). \*\*\**p* =0.0004, \*\**p =*0.0061, and \*\**p* = 0.0052 (day 1) and \**p* =0.0116, \**p =*0.0252, and \**p* = 0.0147 (day 2) for N323Tfs*36, G328Afs*31, and G331Efs*28 mutants, respectively, by two-way ANOVA with Dunnett’s multiple comparisons test. **e,** Quantification of cell survival for GFP-positive cells from day 1 post differentiation split into 3 categories: The range of GFP intensity was 500-140,000. Cells whose GFP intensity was below 45,000, between 45,000 and 90,000, or above 90,000 were grouped into low, medium, or high GFP-expressing cells, respectively. Values represent means ± s.e.m. from n = 3 independent experiments. \**p* =0.0364 and \*\**p* = 0.0088 by one-way ANOVA with Tukey’s multiple comparisons test.

### Alteration or absence of the wild type hnRNPA2 C-terminal sequence, not a neomorphic sequence, is responsible for the cellular phenotypes associated with its frameshift variants

We found it intriguing that all OPMD-associated variants we identified had a +1 frameshift (a deletion of single nucleotide or duplication of two nucleotides) resulting in the same C-terminal rearranged amino acid sequence (Fig. 3b). This rearrangement of the C-terminal amino acid sequence by +1 frameshift introduces several negatively charged amino acids and makes the C-terminal peptide more hydrophobic than the WT peptide (Fig. 5a, b). In hnRNPA1, a homologous protein of hnRNPA2B1 with an M9-NLS, post-translational modifications of residues C-terminal to the conserved PY residues have been shown to inhibit the interaction of hnRNPA1 with Kapβ2 and impair nuclear import of hnRNPA1, indicating that C-terminal flanking regions can regulate PY-NLS activity^25^. Thus, we tested whether the negatively charged anomalous sequence was directly responsible for the phenotypes associated with frameshift variants of hnRNPA2. To this end, we generated two new constructs in which the C-terminal flanking sequence was either deleted (Δ323-341) or +2 frameshifted by a two-base-pair deletion (N323Lfs*31) (Fig. 5b). Interestingly, both Δ323-341 and N323Lfs*31 proteins accumulated in the cytoplasm (Fig. 5c) and caused cell toxicity in differentiated myoblasts at levels comparable to the OPMD-associated frameshift variants (Fig. 5d-e, Supplementary Fig. 4). This result suggests that loss of the wild type C-terminal flanking sequence, rather than addition of a specific, negatively charged C-terminal sequence, is responsible for the observed cellular phenotypes associated with *hnRNPA2B1* frameshift variants.

**Fig. 5:**
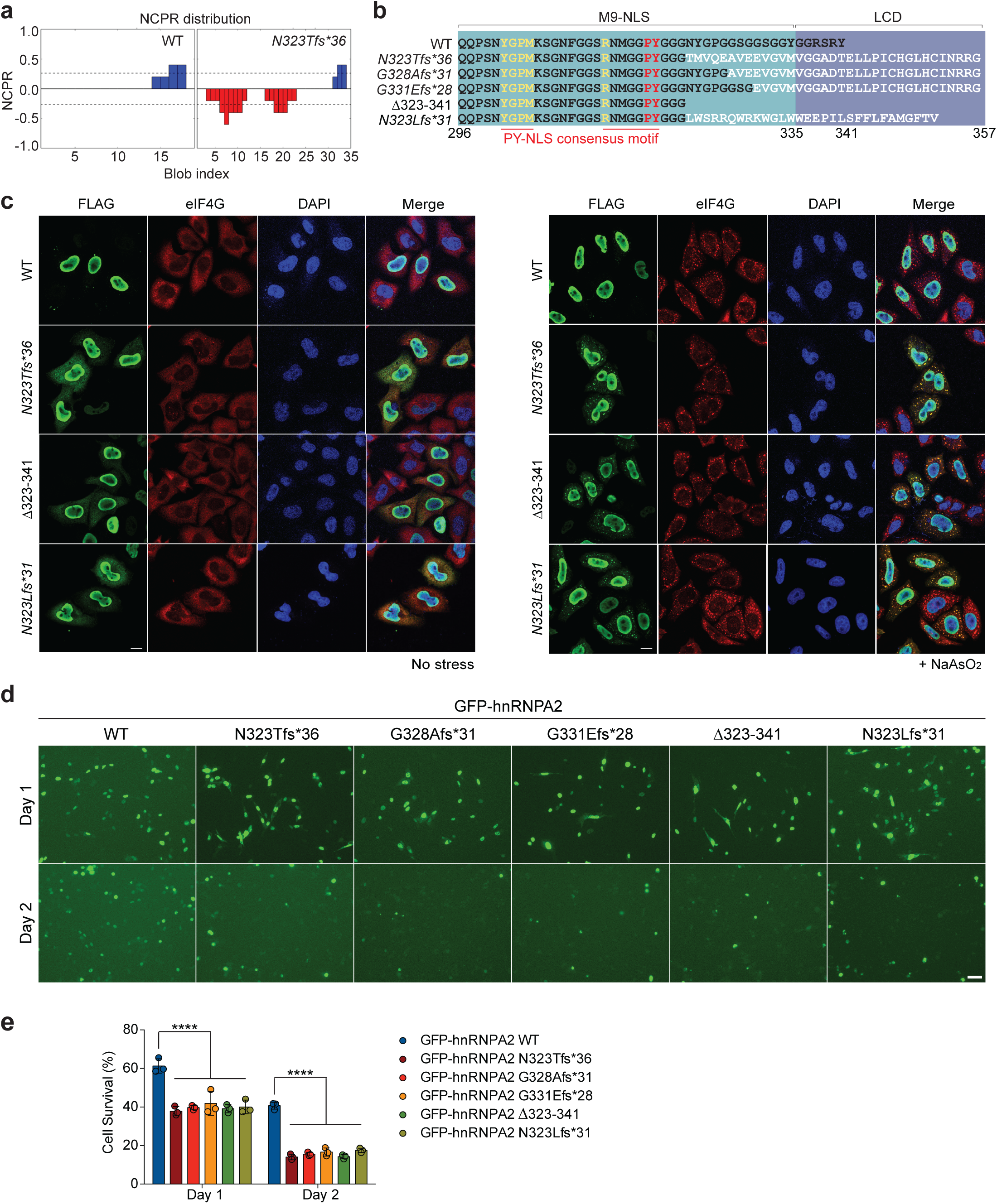
A loss of wild type C-terminal sequence, not a neomorphic sequence, is responsible for phenotypes associated with frameshift variants. **a,** NCPR (net charge per residue; 5-amino acid window) analyses of WT and N323Tfs*36 mutant peptides. **b**. The amino acid sequences of the relevant domains in WT, +1 frameshift mutants identified in OPMD, Δ323-341, and +2 frameshift mutant (N323Lfs*31). **c,** Intracellular localization of FLAG-tagged hnRNPA2 WT, N323Tfs*36, **Δ**323-341, and N323Lfs*31 in the absence (left) or presence (right) of 0.5 mM NaAsO_2_. eIF4G was used as a cytoplasmic and stress granule marker. Scale bar, 10 μm. **d,** Representative images of C2C12 cells one (top) or two (bottom) days after change to differentiation media. Scale bar, 20 μm. **e,** Quantification of GFP-positive cells that were negative for both PI and annexin V. Values represent means ± s.e.m. (n = 3). \*\*\*\**p* < 0.0001 by two-way ANOVA with Dunnett’s multiple comparisons test.

### Frameshift variants cause eye and muscle degeneration in a *Drosophila* model

We next investigated the *in vivo* effects of frameshift variants in a *Drosophila* model system. We generated transgenic *Drosophila* expressing human hnRNPA2 WT, the MSP–associated mutant D290V^6^, or frameshift mutant forms (N323Tfs*36, G328Afs*31, and G331Efs*28) via PhiC31 integrase-mediated site-specific insertion of a single copy of the human *hnRNPA2* gene. We established multiple fly lines per genotype and observed that all lines expressing the N323Tfs*36 mutant and some lines expressing the G328Afs*31 and G331Efs*28 showed low or no protein expression with a GMR-GAL4 driver (Supplementary Fig. 5a). Flies expressing G328Afs*31 and G331Efs*28 at levels comparable to WT protein exhibited mild eye degeneration at 22 °C, whereas flies expressing hnRNPA2 WT or D290V did not display an eye phenotype under the same conditions (Fig. 6a). Increasing expression levels of the transgene by increasing the temperature to 25°C^26^ caused mild eye degeneration in flies expressing hnRNPA2 WT or D290V, severe eye degeneration in flies expressing G331Efs*28, and pupal lethality in flies expressing G328Afs*31 (Fig. 6a). One lethality “escaper” G328Afs*31-expressing fly exhibited severe eye degeneration (Fig. 6a).

**Fig. 6:**
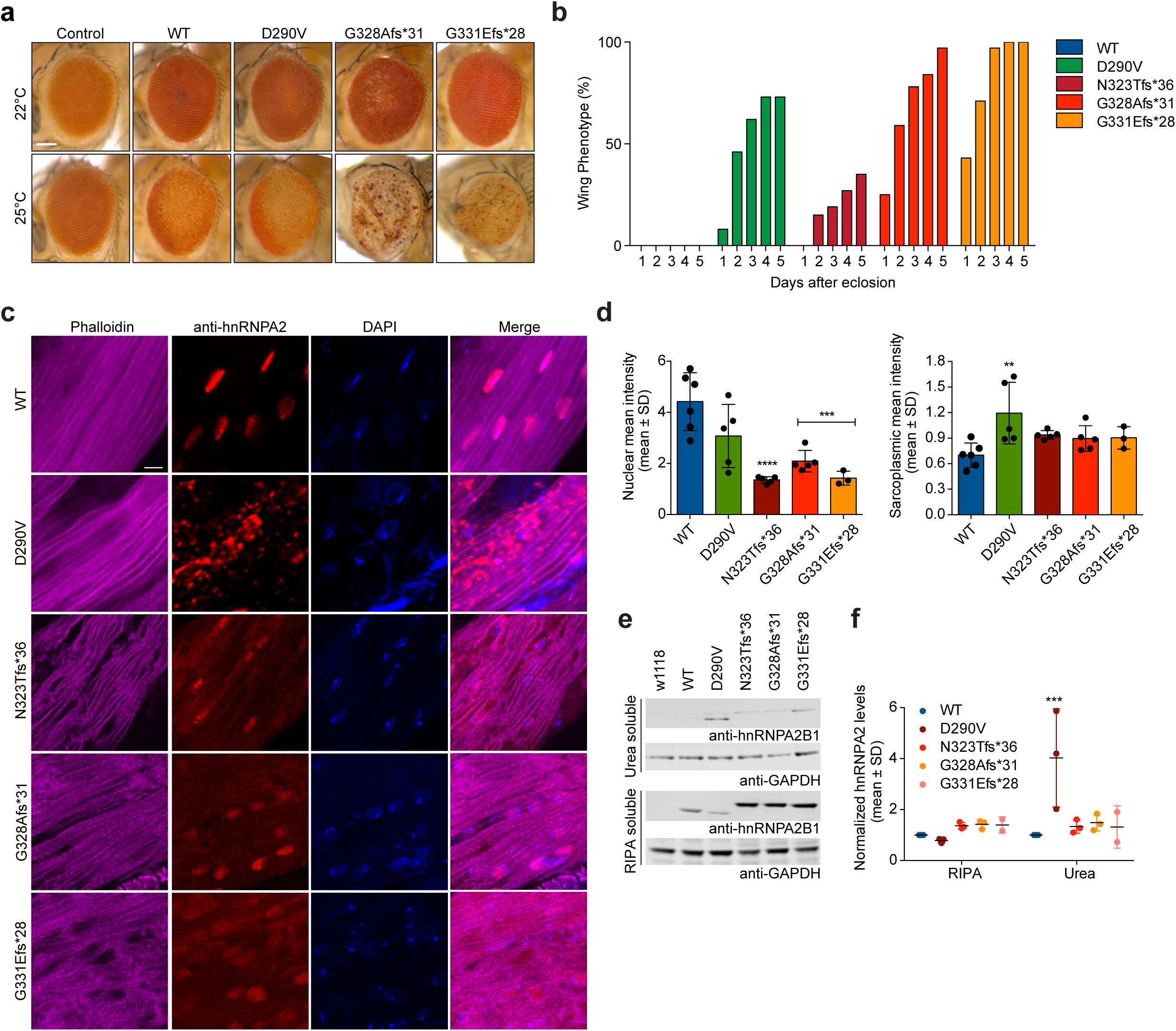
Frameshift variants cause eye and muscle degeneration in a *Drosophila* model. **a,** Expression of frameshift variants in *Drosophila* eye tissue using GMR-GAL4 causes a rough eye phenotype. Scale bar, 100 μm. **b,** Expression of frameshift variants in *Drosophila* muscle using MHC-GAL4 causes an abnormal wing posture phenotype. The percentage of flies with abnormal wing posture is plotted for 5 days after eclosion. n=26, 26, 26, 32, and 35 flies for WT, D290V, N323Tfs*36, G328Afs*31, and G331Efs*28, respectively. **c,** Adult flies expressing hnRNPA2 transgene using MHC-GAL4 were dissected to expose the dorsal longitudinal indirect flight muscle and stained with rhodamine-phalloidin (purple), hnRNPA2 (red), and DAPI (blue). hnRNPA2 WT localized exclusively to nuclei, whereas hnRNPA2 D290V accumulated extensively in cytoplasmic inclusions. Frameshift mutants showed both nuclear staining and diffuse sarcoplasmic accumulation. Scale bar, 5 μm. **d,** Nuclear and sarcoplasmic mean intensity of hnRNPA2 signal in indirect flight muscles. Error bars represent mean ± s.d. \*\**p* < 0.01, \*\*\**p* < 0.001, and *\*\*\*\*p* < 0.0001 compared with WT by two-way ANOVA with Dunnett’s multiple comparisons test. **e,** Thoraces of adult flies expressing hnRNPA2 transgene using MHC-GAL4 were dissected and sequential extractions were performed to examine the solubility profile of hnRNPA2. w1118 flies are included as a non-transgenic control. **f,** Quantification of RIPA soluble and insoluble fraction of hnRNPA2. Data represent mean ± s.d., \*\*\**p* < 0.001 by two-way analysis of variance with Dunnett’s multiple comparisons test.

When expression was driven by the muscle-specific driver MHC-GAL4, we observed robust expression of N323Tfs*36, G328Afs*31, and G331Efs*28 mutant proteins, in contrast with the low expression induced by GMR-GAL4 (Supplementary Fig. 5b). Although a single copy of the *hnRNPA2* gene was inserted in all lines, the levels of protein expression were consistently modestly higher in the N323Tfs*36, G328Afs*31, and G331Efs*28 flies compared with the hnRNPA2 WT and D290V flies, suggesting that the frameshift mutations may increase the stability of hnRNPA2 protein in fly muscles (Supplementary Fig. 5b). Expression of the D290V, N323Tfs*36, G328Afs*31, and G331Efs*28 mutants caused age-dependent defects in indirect flight muscles, resulting in a greater prevalence of abnormal wing position compared with flies expressing hnRNPA2 WT (Fig. 6b). Expression of the D290V, N323Tfs*36, G328Afs*31, and G331Efs*28 mutants all caused severe muscle degeneration (Fig. 6c). Immunohistochemical analysis of indirect flight muscles showed that hnRNPA2 WT localized appropriately to nuclei, whereas the D290V mutant largely accumulated in cytoplasmic inclusions, as previously reported (Fig. 6c)^6,27^. In contrast, N323Tfs*36, G328Afs*31, and G331Efs*28 proteins exhibited both nuclear and diffuse cytoplasmic localization and did not form cytoplasmic inclusions (Fig. 6c-d).

Cytoplasmic aggregate formation was associated with the solubility of hnRNPA2 protein as expressed in thoracic tissue from *Drosophila*. Specifically, D290V mutant protein was largely found in the detergent-insoluble (urea) fraction, as previously reported^6^, whereas hnRNPA2 WT, N323Tfs*36, G328Afs*31, and G331Efs*28 proteins were found mostly in the detergent-soluble (RIPA) fraction (Fig. 6e,f). Consistent with these findings, N323Tfs*36 and G328Afs*31 exhibited decelerated fibrillization kinetics at the pure protein level (Supplementary Fig. 6). Thus, purified recombinant hnRNPA2 WT readily formed fibrils, and hnRNPA2 D290V assembled into fibrils at a faster rate^6,12^ (Supplementary Fig. 6a). In contrast, the N323Tfs*36 mutant formed very few fibrils over 24 h, whereas the G328Afs*31 mutant fibrillized only after a long lag phase (Supplementary Fig. 6a,b). Thus, the frameshift mutations reduce the intrinsic propensity for hnRNPA2 to fibrillize. These findings strongly suggest that the pathogenic mechanism of the frameshift variants differs from that of D290V and likely involves reduced efficiency of nuclear import.

## Discussion

Here we describe a distinct phenotype of early-onset myopathy associated with heterozygous *hnRNPA2B1* frameshift variants clinically manifesting with ptosis, ophthalmoplegia, dysphagia, and variable degrees of respiratory insufficiency and progressive muscle weakness. With the notable exception of the very early disease onset and rapid progression in our cohort, the overall clinical presentation overlapped with classical OPMD, albeit with a few characteristic differences. The uniform absence of bone, cognitive and motor neuron involvement in our patients distinguishes our cohort from the previously described MSP phenotype associated with a p.D290V *hnRNPA2* variant^6^. In addition, ptosis, ophthalmoplegia, and dysphagia were absent in the reported *hnRNPA2* missense (D290V) family, whereas these were consistent phenotypic features within our patient cohort, further differentiating these distinct *hnRNPA2B1*–related phenotypes.

Since the identification of the first *hnRNPA2B1* MSP family, screening of MSP and large ALS patient cohorts has shown that *hnRNPA2B1* variants are a rare cause of sporadic and familial motor neuron disease^28–30^; however, its role in myopathies remains elusive. Moreover, the recent identification of the p.P298L missense *hnRNPA2B1* variant in a family with isolated Paget’s disease of the bone without multisystem involvement highlights the divergent phenotypes that may arise from the various *hnRNPA2B1* disease variants^13^. In contrast with the previously reported variants, the *hnRNPA2B1* variants reported here are all specific frameshift variants that cluster in the highly conserved PrLD of the protein and escape RNA quality control degradation (Supplementary Fig. 2). These frameshift *hnRNPA2B1* variants alter its C-terminal M9-NLS amino acid sequence while sparing the immediately upstream PY-NLS residues. Further investigation of the sequence features that contribute to frameshift variants associated phenotypes indicates that loss of the wild type C-terminal sequence, rather than gain of an additional amino acid sequence in frameshift variants, is responsible for the cellular phenotypes we observed (Fig. 5, Supplementary Fig. 4). Thus, we propose that the C-terminal flanking sequence plays an important role in hnRNPA2B1 functions and its loss may be an underlying molecular mechanism for this class of frameshift *hnRNPA2B1* variants, which consistently manifest as a newly described early-onset OPMD-like phenotype. Whether the same OPMD-like phenotype will be observed with truncation or other alterations of the *hnRNPA2B1* C-terminal flanking sequence in patients remains unclear.

Nuclear-import receptors regulate the LLPS, fibrillization, and nuclear import of PrLD-containing RBPs^31–33^. While MSP/PDB-associated missense *hnRNPA2* variants directly promote fibrillization and the formation of cytoplasmic protein aggregates, frameshift *hnRNPA2B1* variants show reduced propensity for fibrillization (Supplementary Fig. 6), reduced affinity for Kapβ2 (Fig. 3), and impaired nuclear import (Fig. 3). This contrast in protein aggregation and subcellular distribution between missense and frameshift *hnRNPA2B1* variants was noted in cells^6^ (Fig. 3c-f) and *Drosophila* indirect flight muscle (Fig. 6c-f), and hence our experimental models parallel the divergent phenotypes associated with the distinct variant classes.

All the *hnRNPA2B1* frameshift variants reported here were found in heterozygosity and appear to act in a dominant manner. This observation raises the possibility of loss of function (i.e., haploinsufficiency), gain of function, or dominant-negative pathomechanisms. In our studies, high expression levels of WT and frameshift hnRNPA2 had similar effects on cell survival as the expression of frameshift hnRNPA2B1 protein (Fig. 4e), hinting at an underlying gain-of-function mechanism. This effect may arise in part due to increased stability of the mutant protein as suggested in the *Drosophila* models (Supplementary Fig. 5b). Unlike the missense p.D290V variant that promotes fibrillization and appears to be almost exclusively cytoplasmic in distribution (Fig 6c), frameshift hnRNPA2B1 variants have a reduced propensity for fibrillization compared to WT protein (Supplementary Fig. 6) and display both nuclear and cytoplasmic localization in cells and *Drosophila* muscle (Fig. 6c), which argues against a similar dominant-negative effect. Dosage sensitivity, related to hnRNPA2B1’s intranuclear function, remains another plausible pathomechanism and is supported by a relative paucity of loss-of-function alleles in the gnomAD database (pLI = 1, observed-to-expected ratio=0.05)^15^. These two mechanisms are not mutually exclusive and may both be relevant for the pathophysiology of the disease: Since the altered distribution of the frameshift hnRNPA2B1 variant simultaneously imposes its relative abundance in the cytoplasm and its relative deficiency in the nucleus, the downstream cellular effects of this class of variants is likely complex, multifold, and variable in different cell types.

Muscle pathologic features in our *hnRNPA2B1* cohort were overall consistent with a chronic myopathic process, with notable histological findings of rimmed autophagic vacuoles and cytoplasmic and intranuclear tubulofilamentous inclusions. These non-specific findings have been reported in several myopathies, including *hnRNPA2B1* missense variant MSP, inclusion body myositis^34^, *MYH2-*related inclusion body myopathy^35^, *hnRNPA1*-associated inclusion body myopathy^36^, and OPMD^37^, although the intranuclear tubulofilamentous inclusions we noted lacked OPMD-associated palisading morphology. The similarity in muscle pathological features of *hnRNPA2B1-*associated MSP and our newly described OPMD-like disease, despite their underlying genetic divergence and stark contrast in promoting vs decelerating protein fibrillization, may appear paradoxical. However, in human pathological studies of ALS/FTD brain samples, which at late stages are also characterized by presence of TDP-43 cytoplasmic aggregates, loss of nuclear TDP-43 staining precedes the development of cytoplasmic inclusions by several years^38^. Thus, it is possible that the hnRNPA2B1 aggregates in our patient biopsies are the end-stage epiphenomenon of the disease process and are preceded by abnormal nucleocytoplasmic transport dynamics of frameshift variant hnRNPA2B1.

Among the myopathies, emerging data suggest a specific role for PrLD-containing RBPs such as TDP-43 and hnRNPA2B1 in regenerating muscle, which presumably act to stabilize large muscle-specific transcripts (e.g., TTN, NEB) and facilitate pre-mRNA splicing^39^. In particular, wild-type hnRNPA2B1 retains its exclusive nuclear localization during muscle regeneration^39^. Consistent with their inefficient nuclear import, where it would exert its normal biological function in muscle, hnRNPA2B1 frameshift variants in our study resulted in cell toxicity in differentiating C2C12 myoblasts (Fig. 4) and disrupted the development of flight muscle in *Drosophila* (Fig. 5), confirming an essential role for nuclear hnRNPA2B1 in developing and regenerating muscle. Thus, with ongoing use and stress of the muscle, impaired hnRNPA2B1 nuclear import may be inadequate to maintain muscle homeostasis over time, clinically manifesting with the progressive muscle weakness seen in our patients. The phenotypic overlap of our patients with OPMD is also of great interest, specifically as OPMD is caused by poly-alanine expansions in *PABPN1*, whose normal splicing in the nucleus is partially regulated by hnRNPA2B1^40^. Taken together, these findings suggest the possibility of concurrent nuclear loss-of-function and cytoplasmic gain-of-function mechanisms. Thus, future studies should focus on the role of RBPs such as hnRNPA2B1 in specific cell types, specific subcellular compartments and in response to different physiologic states to identify their respective contributions to pathogenic processes and subsequent clinical manifestation.

Our data expand the clinical spectrum of *hnRNPA2B1* variants from MSP to now also include a distinct early-onset OPMD-like phenotype. It is not surprising that *hnRNPA2B1* variants are involved in such broad disease phenotypes, given the fundamental role of hnRNPA2B1 in mRNA processing, transport, and stability. Understanding how these seemingly divergent phenotypes emerge from common molecular and cellular events will uncover fundamental regulatory mechanisms of RBPs with prion-like domains that have applicability in a broad array of neurodegenerative diseases.

## Data Availability

All data generated or analysed during this study are included in this published article (and its supplementary information files).

## Acknowledgements

We thank the patients and their families for participating in our research study. We thank Christopher Mendoza, and Gilberto (“Mike”) Averion for their help in clinic. We also thank the NIH Intramural Sequencing Center staff and Daniel MacArthur and Fengmei Zhao (Analytic and Translational Genetics Unit at Massachusetts General Hospital in collaboration with the Broad Institute) for their help with the exome analysis. We thank Darren Chambers and Lucy Feng for technical assistance in processing muscle pathology specimens.

Work in C.G. B.’s laboratory is supported by intramural funds from the NIH National Institute of Neurological Disorders and Stroke. Exome sequencing was in part funded through the Clinical Center Genomics Opportunity, which is sponsored by the National Human Genome Research Institute, the NIH Deputy Director for Intramural Research, and the NIH Clinical Center. J.P.T. is supported by the Howard Hughes Medical Institute, R35NS097974, and the ALS Association (18-IIA-419). This study was supported in part by Intramural Research Grant (2-5 and 29-4 to IN; 2-5 and 30-9 to AI) for Neurological and Psychiatric Disorders of NCNP and by AMED under Grant Nos. 20ek0109490h0001 and JP19ek0109285h0003 (to IN) and Joint Usage and Joint Research Programs, the Institute of Advanced Medical Sciences, Tokushima University (2020, 2A19 to AI). The work in F.M. group was supported by the National Institute for Health Research Biomedical Research Centre at Great Ormond Street Hospital for Children NHS Foundation Trust and University College London and the MRC Centre for Neuromuscular Diseases Biobank, and by the HSS England Diagnostic and Advisory Service for Congenital Myopathies and Congenital Muscular Dystrophies in London, UK for their financial support to the DNC Muscle Pathology Service. F.M. was also funded by the European Community’s Seventh Framework Programme (FP7/2007-2013) under grant agreement n° 2012-305121 “Integrated European –omics research project for diagnosis and therapy in rare neuromuscular and neurodegenerative diseases (NEUROMICS)”. E.P. and L.B. are members of the European Reference Network for Neuromuscular Diseases – Project ID N° 870177 and acknowledge support from the Telethon Network of Genetic BioBank (GTB12001D) and EuroBioBank network. The work in J.L. group is supported by the France Génomique National infrastructure and by the Fondation Maladies Rares within the frame of the “Myocapture” sequencing project, and by Association Française contre les Myopathies (22734). C.E.F and F.L.R. are funded by Cambridge NIHR Biomedical Research Centre and the Rosetree Foundation. L.G. was supported by an Ellison Medical Foundation/American Federation for Aging Research fellowship, Alzheimer’s Association Research fellowship, and a Target ALS Springboard Fellowship. C.M.F. was supported by NIH grants T32GM008275 and F31NS111870. A.F.F. was supported by NIH grants T32AG00255 and F31NS087676. J.S. was supported by Target ALS, Packard Foundation for ALS research, ALS Association, The G. Harold and Leila Y. Mathers Charitable Foundation, and NIH grant R01GM099836. MYOSEQ was funded by Sanofi Genzyme, Ultragenyx, LGMD2I Research Fund, Samantha J. Brazzo Foundation, LGMD2D Foundation and Kurt+Peter Foundation, Muscular Dystrophy UK, and Coalition to Cure Calpain 3. Analysis was provided by the Broad Institute of MIT and Harvard Center for Mendelian Genomics (Broad CMG) and was funded by the National Human Genome Research Institute, the National Eye Institute, and the National Heart, Lung, and Blood Institute grant UM1 HG008900, and in part by National Human Genome Research Institute grant R01 HG009141.

## Supplementary Figure Legends

**Supplementary Fig. 1.**
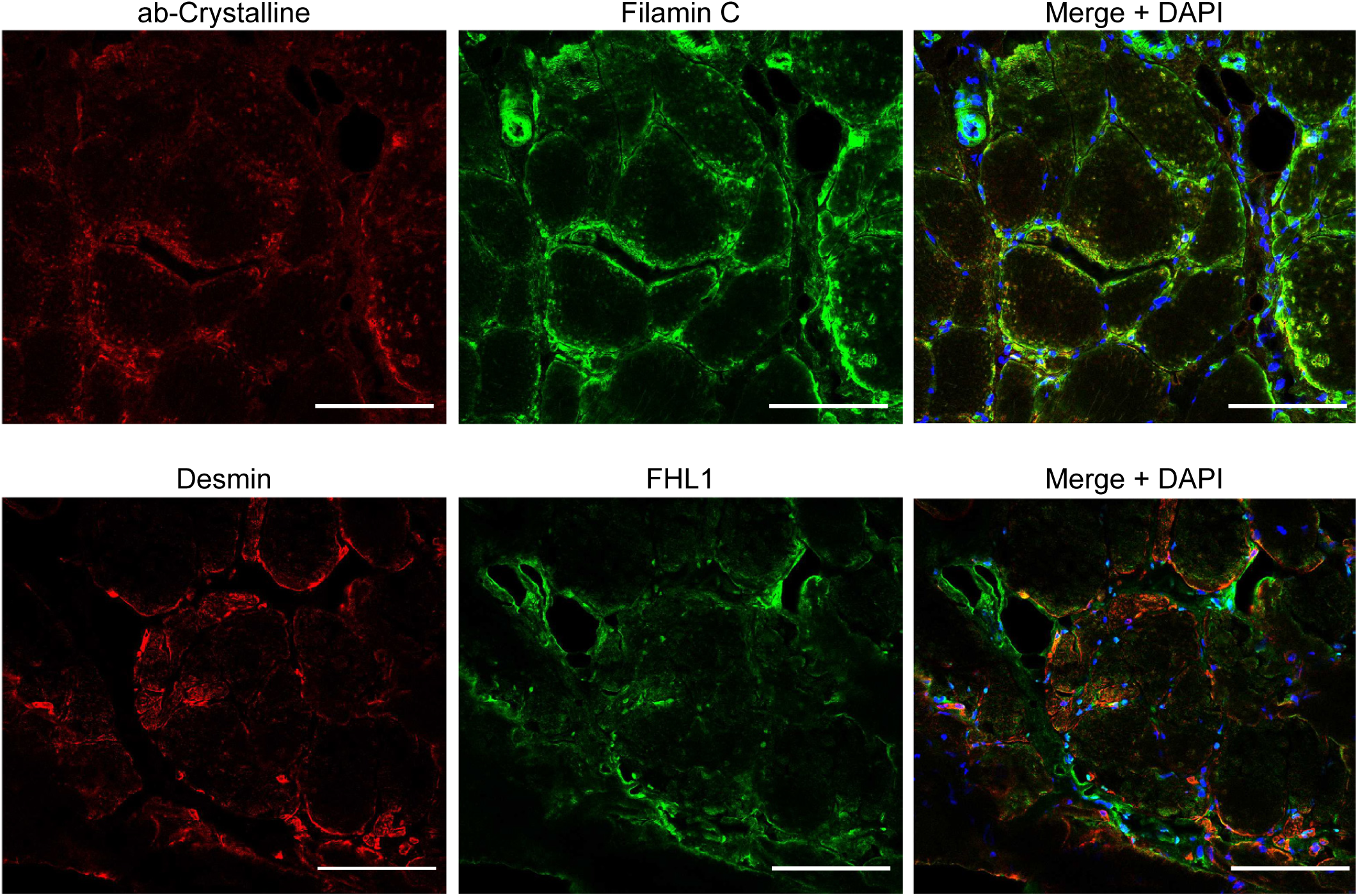
Cytoplasmic myofiber inclusions do not contain myofibrillar proteins. Immunostaining of patient 1 muscle biopsy with αβ-crystalline, filamin C, desmin and FHL1 showing absence of cytoplasmic or perinuclear inclusions or protein aggregates that contain these Z-disk associated myofibrillar proteins.

**Supplementary Fig. 2.**
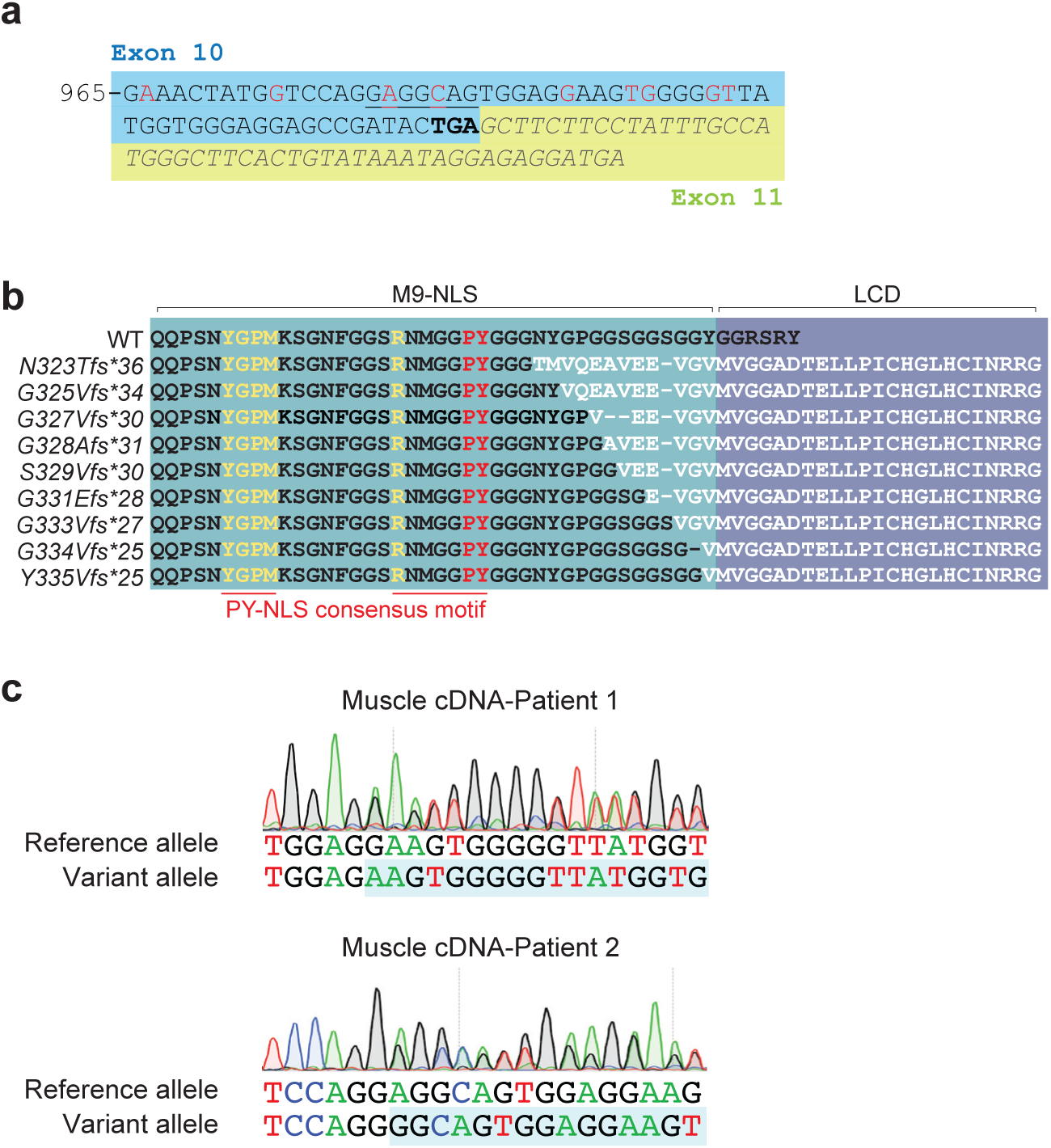
Frameshift *hnRNPA2B1* variant transcripts are stable in muscle tissue and cause rearrangement of amino acid sequence in the C terminus. **a,** cDNA sequence of *hnRNPA2* (NM_002137) in exon 10 and exon 11. Red nucleotides indicate nucleotides deleted in patients; underlined nucleotides indicate nucleotides deleted in patient 8. The original stop codon (TGA) is indicated in bold. **b**, Amino acid sequences in WT and each frameshift mutant. The consensus PY-NLS motif in M9-NLS is underlined in red on bottom row. **c**, RNA was extracted from muscle tissue of patients 1 (c.992delG) and 2 (c.981delA). After reverse transcriptase reaction, the cDNA was amplified by PCR and sequenced. The intensity of the chromatogram peaks corresponding to the reference and frameshift alleles appear nearly equal, suggesting little to no degradation of the frameshift transcript in muscle tissue.

**Supplementary Fig. 3.**
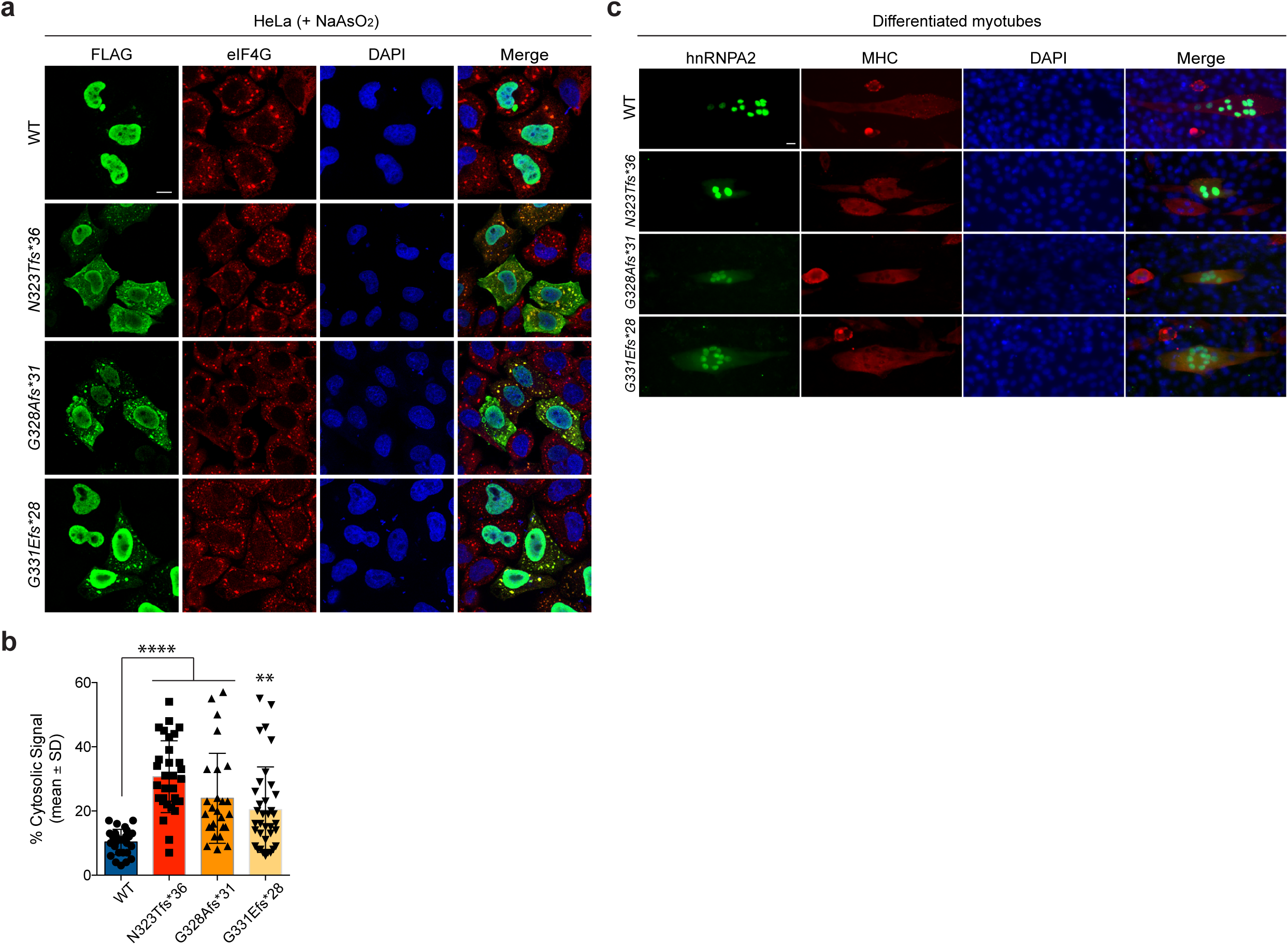
Frameshift variants enhance hnRNPA2B1 cytoplasmic localization and recruitment to RNP granules. **a,** Intracellular localization of FLAG-tagged hnRNPA2 WT, N323Tfs*36, G328Afs*31, and G331Efs*28 mutants upon 30 min 0.5 mM NaAsO_2_ stress in HeLa cells. **b,** Quantification of hnRNPA2 cytosolic signal intensity in cells as shown in (a). An interleaved scatter plot with individual data points is shown; error bars represent mean ± s.d. For WT, N323Tfs*36, G328Afs*31, and G331Efs*28 mutants, n=27, 31, 26, and 36 cells, respectively. *\*\*\*\*p* < 0.0001 by two-way ANOVA with Dunnett’s multiple comparisons test. **c,** Intracellular localization of hnRNPA2 WT, N323Tfs*36, G328Afs*31, and G331Efs*28 mutants in differentiated C2C12 myotubes.

**Supplementary Fig. 4.**
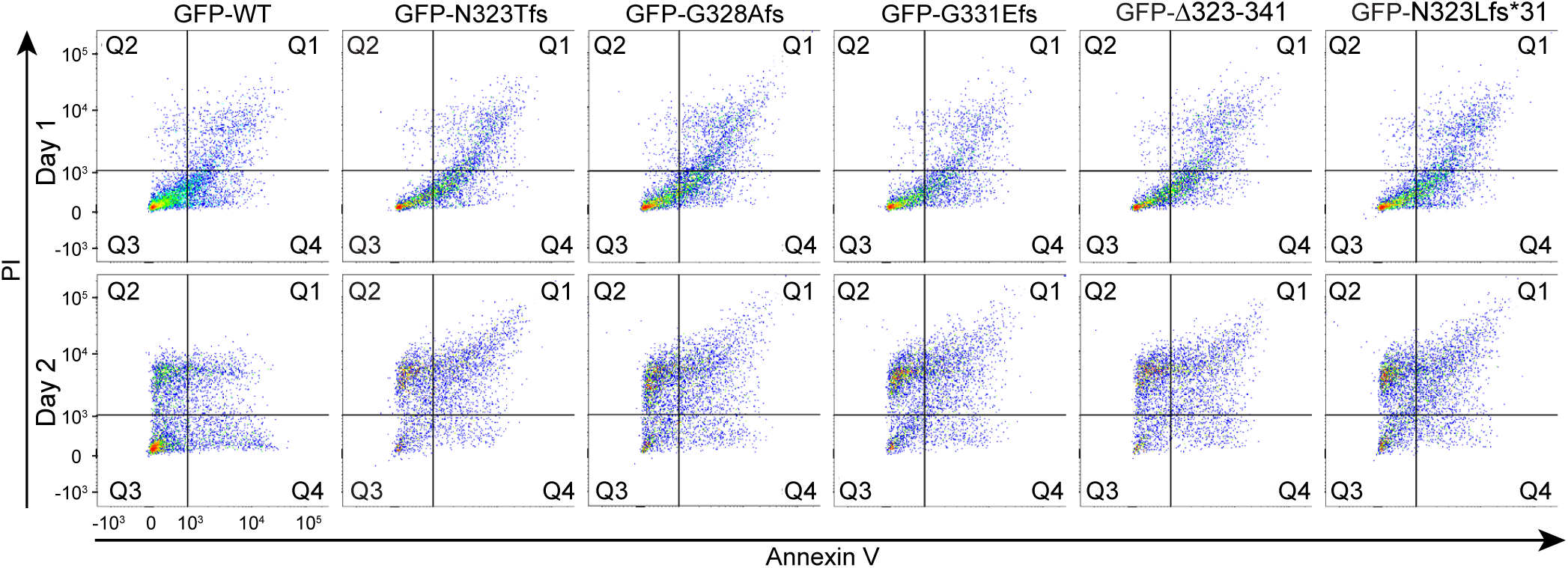
Alteration or absence of the wild type hnRNPA2 C-terminal sequence, not a neomorphic sequence, is responsible for apoptotic cell death in differentiating cells. Representative scatter plots of GFP-positive C2C12 cells stained with propidium iodide (PI) and annexin V, one (top) or two (bottom) days after change to differentiation media.

**Supplementary Fig. 5.**
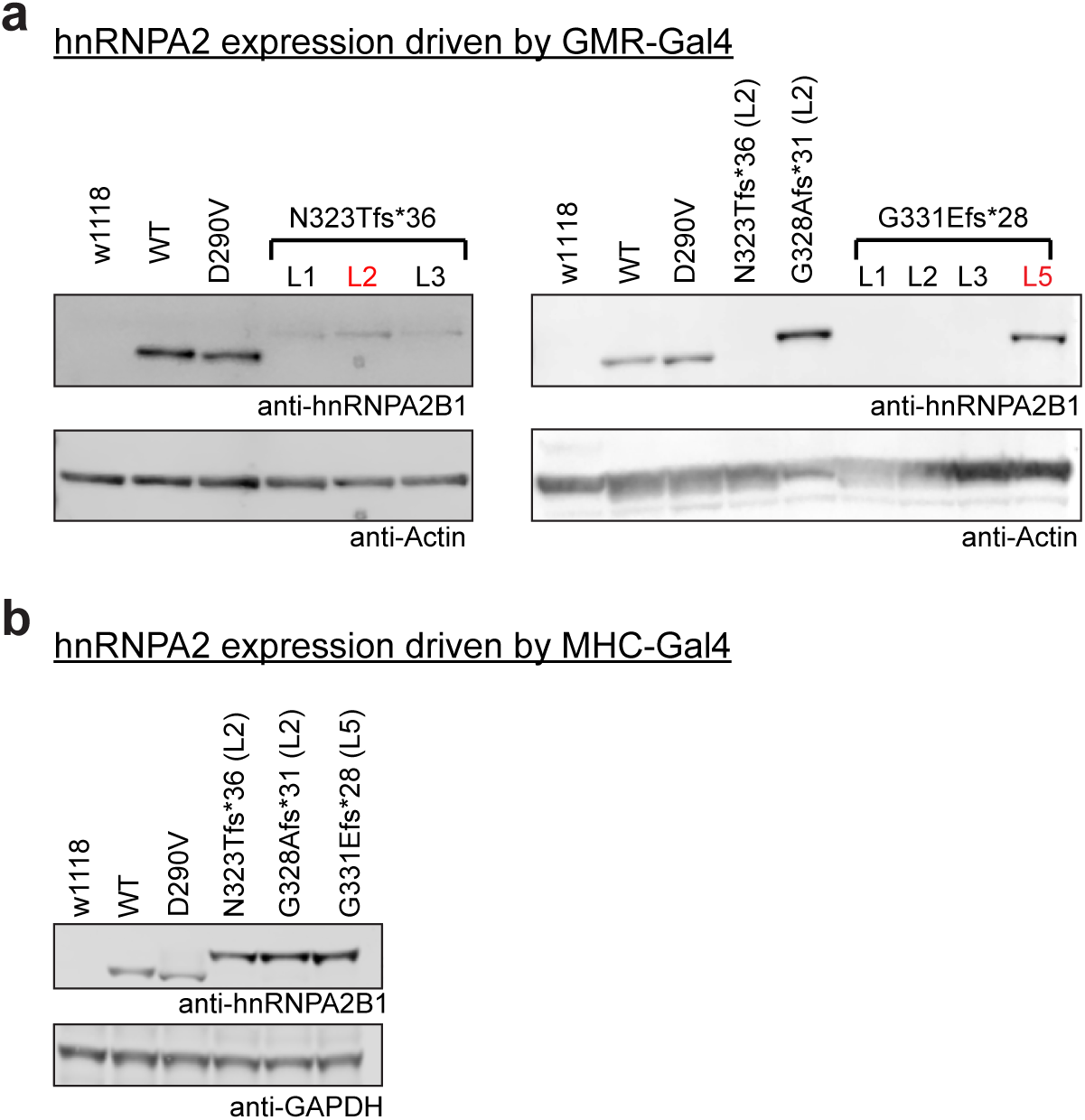
Expression of frameshift variants in a *Drosophila* model. **a,** hnRNPA2 expression driven by GMR-GAL4 in transgenic flies. Lysates were prepared from heads of adult flies and analyzed by Western blot using an antibody against hnRNPA2B1. Actin was used as a loading control. Lines (L) selected for further experiments are indicated in red. **b,** hnRNPA2 expression driven by MHC-GAL4 in transgenic flies. Lysates were prepared from thoraces of adult flies and analyzed by Western blot using an antibody against hnRNPA2B1. GAPDH was used as a loading control.

**Supplementary Fig. 6.**
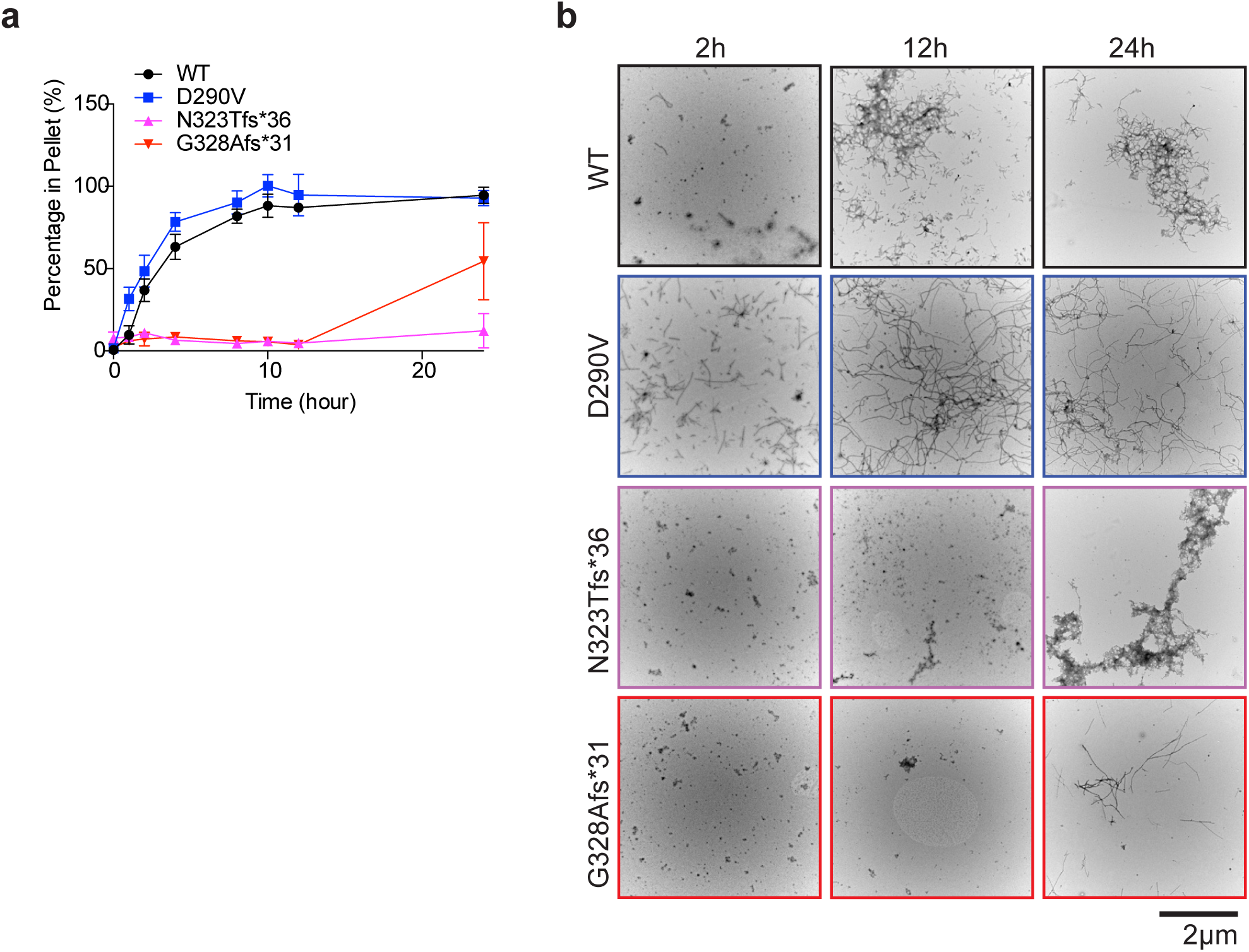
Frameshift variants reduce the intrinsic ability of hnRNPA2 to fibrillize. **a,** Full-length purified hnRNPA2 WT, D290V, N323Tfs*36, or G328Afs*31 (5µM) were incubated at 25°C with agitation for 0-24 h. The amount of aggregated hnRNPA2 was determined by sedimentation analysis at indicated times. Values represent means ± s.e.m. (n=3). **b,** Electron micrographs of hnRNPA2 fibrillization reactions after 2, 12 and 24 h at 25°C. Fibrils were scarce for WT after 2 h, abundant for D290V after 2 h, and absent for N323Tfs*36 and G328fs*31 after 2 h. Fibrils remained absent after 12 h for hnRNPA2 N323Tfs*36 and G328fs*31, but were abundant for D290V and WT. After 24 h, all hnRNPA2 variants formed fibrils, but fibrils were less abundant for N323Tfs*36 and G328fs*31. Scale bar, 2 μm.

## Online Methods

### Patient recruitment and sample collection

Patients were recruited through local neurology and genetics clinics and clinical information was obtained based on the local standard clinical care. DNA and tissues (e.g., muscle, skin biopsy) and medical records were obtained based on standard procedures. For research studies, written informed consent and age-appropriate assent was obtained from all participants. Ethical approval was obtained from the NIH, National Institute of Neurological Disorders and Stroke (NINDS), Institutional Review Broad (Protocol 12-N-0095), National Center of Neurology and Psychiatry (Protocol A2019-123), University of Strasburg (Protocol DC-2012-1693), Cambridge South, UK Research Ethics Committee (approval 13/EE/0325), Health Research Authority, NRES Committee East of England – Hatfield (REC 13/EE/0398; REC 06/Q0406/33) and National Research Ethics Service (NRES) Committee North East–Newcastle & North Tyneside 1 (reference 08/H0906/28).

### Exome sequencing

Quartet exome sequencing in family 1 was performed through the NIH Intramural Sequencing Center (NISC) using the Illumina TruSeq Exome Enrichment Kit and Illumina HiSeq 2500 sequencing instruments. Variants were analyzed using *Seqr* (Center for Mendelian Genomics, Broad Institute). Trio exome sequencing in family 2 was performed at GeneDX with exon targets isolated by capture using the Agilent SureSelect Human All Exon V4 (50 Mb) kit or the Clinical Research Exome (Agilent Technologies). The sequencing methodology and variant interpretation protocol has been previously described^41^.

Patient 3 was sequenced as a singleton as part of the MYO-SEQ project^42^. Exome sequencing was performed by the Genomics Platform at the Broad Institute of MIT and Harvard, Cambridge, USA. Libraries were created with an Illumina exome capture (38 Mb target) and sequenced with a mean target coverage of >80x. Exome sequencing data were analysed on *seqr* (https://seqr.broadinstitute.org/).

For family 4, exome sequencing of patients 4 and 5 was undertaken using Agilent SureSelect Human All Exon 50Mb capture kit followed by sequencing on a 5500XL SOLiD sequencer. The affected mother and unaffected father were sequenced using Nextera Rapid Capture Expanded Exome for target selection followed by sequencing on an Illumina HiSeq2000. Genomic data were processed as previously described^43^. Confirmation of variants and segregation was performed by Sanger sequencing.

WES analysis in proband (P6) from Family 5 was performed by deCODE genetics, Iceland. The alignment to the human reference genome (hg19) and the variant calling was done by deCODE genetics, Iceland. Data analysis was carried out using Clinical Sequence Miner platform, NextCODE genetics.

Trio exome sequencing in family 6 was performed at the Centre National de Recherche en Génomique Humaine (Evry, France) using the Agilent SureSelect Human All Exon V4. Sequence analysis and variant interpretation was performed as described^44^.

For family 7, the patient underwent standard clinical next generation sequencing panels that included LGMD and congenital myopathy genes. Due to the remarkably similar phenotype to the other patients in this cohort, direct sanger sequencing of the terminal exon of *hnRNPA2B1* was pursued and identified the *de novo* variant.

For family 8, the patient’s DNA was analyzed using the Illumina Trusight ONE Expanded kit on an Illumina NextSeq 550 sequencer as described^45^. Segregation analysis was performed by Sanger sequencing.

For family 9, whole genome sequencing in patient 10 and her parents was performed using the Illumina HiSeq X Ten platform and variant calling and interpretation were performed as previously described^46^. Confirmation of variants was performed by Sanger sequencing.

For family 10, exome sequencing in patient 11 was performed at Beijing Genomics Institute (BGI) using the DNBSEQ. Exome sequencing and variant calling were performed as previously described^47^. Confirmation of variants was performed by Sanger sequencing.

### Muscle histology, immunofluorescence and confocal microscopy

Clinical muscle biopsy slides were obtained and reviewed. These included hematoxylin and eosin, modified Gömöri trichrome, NADH and other histochemical stains as well as electron microscopy images when available. For immunofluorescence and confocal microscopy, frozen muscle biopsy tissues were cryo-sectioned (10 µm), fixed (100% acetone, −20 °C for 10 min) and blocked and permeabilized in 5% normal goat serum (Sigma) with 0.5% Triton X in PBS for 1 h at room temperature. Primary antibody incubation was performed overnight at 4 °C as follows: hnRNPA2B1 (Santa Cruz, sc-32316; mouse, 1:200), TDP-43 (Proteintech, 10782-2-AP; rabbit, 1:200), ubiquitin (Stressgen, SPA-200; rabbit, 1:200), ubiquilin-2 (Abcam, Ab190283; mouse IgG1, 1:500), TIA1 (Abcam, Ab140595; rabbit, 1:100), P62/SQSTM1 (Santa Cruz, sc-28359; mouse IgG1, 1:250). Secondary antibody incubation was performed for 1 h at room temperature. Sections were then washed in PBS, the nuclei were stained with DAPI, and sections were mounted and coverslipped. Z-stack images were obtained using a Leica TCS SP5 II confocal microscope.

### Muscle MRI

Muscle MRI was performed using conventional T1-weighted spin echo and short tau inversion recovery (STIR) of the lower extremities on different scanners at different centers.

### Plasmid constructs

cDNA containing frameshift mutations of hnRNPA2 were synthesized by GenScript. For mammalian expression, FLAG-tagged WT, D290V, N323Tfs, G328Afs, G331Efs, **Δ**323-341, and N323Lfs*31 were cloned into pCAGGS vector at *SacI* and *SbfI* sites. GFP-tagged WT, D290V, N323Tfs, G328Afs, G331Efs, **Δ**323-341, and N323Lfs*31 were cloned into pEGFP-C1 vector at *BsrGI* and *XhoI* sites. For transgenic *Drosophila*, mutant hnRNPA2 cDNAs were subcloned into the pUASTattB vector using *EcoRI* and *XhoI*. For bacterial expression, codon optimized cDNA containing frameshift mutations of hnRNPA2 were synthesized and subcloned into the pGST-Duet vector using *BamHI* and *EcoRI* by GenScript. All clones were verified by restriction enzyme digestion and sequence analysis.

### Cell culture, transfection and immunofluorescence

HEK293T and HeLa cells were grown in Dulbecco’s modified Eagle’s medium (DMEM) supplemented with 10% fetal bovine serum (FBS), 1% penicillin/streptomycin and 1% L-glutamate. Cells were transfected using FuGene 6 (Promega) according to the manufacturer’s instructions. For immunofluorescence, HeLa cells were seeded on 8-well glass slides (Millipore) and transfected with FLAG-hnRNPA2 WT, FLAG-hnRNPA2 N323Tfs, FLAG-hnRNPA2 G328Afs, FLAG-hnRNPA2 G331Efs, FLAG-hnRNPA2 **Δ**323-341, or FLAG-hnRNPA2 N323Lfs*31 construct. 24 h post transfection, cells were stressed with 500 μM sodium arsenite (Sigma-Aldrich) for indicated times. Cells were then fixed with 4% paraformaldehyde (Electron Microscopy Sciences), permeabilized with 0.5% Triton X-100 and blocked in 3% bovine serum albumin. Primary antibodies used were mouse monoclonal anti-FLAG (M2, F1804; Sigma), rabbit polyclonal anti-eIF4G (H-300, sc-11373; Santa Cruz Biotechnology), and mouse monoclonal anti-hnRNPA2B1 (EF-67, sc-53531; Santa Cruz Biotechnology) antibodies. For visualization, the appropriate host-specific Alexa Fluor 488, 555 or 647 (Molecular Probes) secondary antibody was used. Slides were mounted using Prolong Gold Antifade Reagent with DAPI (Life Technologies). Images were captured using a Leica TCS SP8 STED 3X confocal microscope (Leica Biosystems) with a 63x objective.

### Western blot analysis

Cell lysates were prepared by lysing cells in 1× lysis buffer (150 mM NaCl, 25 mM Tris-HCl pH 7.5, 1 mM EDTA, 5% glycerol and 1% NP-40) with Complete Protease Inhibitor Cocktail (Clontech Laboratories). Samples were resolved by electrophoresis on NuPAGE Novex 4–12% Bis-Tris gels (Invitrogen). Protein bands were visualized using the Odyssey system (LI-COR) and Image Studio (LI-COR). Primary antibodies used were mouse monoclonal anti-FLAG (M2, F1804; Sigma), rabbit polyclonal anti-FLAG (F7425; Sigma), rabbit polyclonal anti-GFP (FL, sc-8334; Santa Cruz Biotechnology), rabbit polyclonal anti-βTubulin (H-235, sc-9104; Santa Cruz Biotechnology), rabbit-polyclonal anti-hnRNPA2B1 (HPA001666, Sigma), rabbit-polyclonal anti-hnRNPA2B1 (NBP2-56497; NOVUS Biologicals), and mouse monoclonal anti-GAPDH antibodies (6C5, sc-32233; Santa Cruz Biotechnology). Blots were subsequently incubated with IRDye fluorescence-labeled secondary antibodies (LI-COR) and protein bands were visualized using the Odyssey Fc system (LI-COR) and Image Studio (LI-COR).

### Cell toxicity

C2C12 cells were grown in Dulbecco’s modified Eagle’s medium (DMEM) supplemented with 10% fetal bovine serum (FBS), 1% penicillin/streptomycin, and 1% L-glutamate. Cells were transfected using Lipofectamine 300 reagent (ThermoFisher Scientific) according to the manufacturer’s instructions. C2C12 cells were plated in 6-well dishes (Corning) and transfected with GFP-, GFP-hnRNPA2 WT, GFP-hnRNPA2 N323Tfs, GFP-hnRNPA2 G328Afs, GFP-hnRNPA2 G331Efs, GFP-hnRNPA2 **Δ**323-341, or GFP-hnRNPA2 N323Lfs*31 construct. constructs. 24 h post transfection, the media was changed to differentiation media (DMEM supplemented with 0.5% FBS, 1% penicillin/streptomycin, and 1% L-glutamate) and counted as day 0. After 1 or 2 days, the GFP signal of the cells were imaged with an EVOS microscope. Apoptotic cells were determined by staining with Annexin-V-APC and propidium iodide (BD Biosciences) and measured by flow cytometry. Data was analyzed using FlowJo_v10.6.1.

### Protein purification

GST-tagged Kapβ2 WT protein was purified as described previously^48^ with modifications. Briefly, recombinant protein was expressed in BL21 (DE3) *Escherichia coli* cells by induction with 1 mM isopropyl-β-D-thiogalactoside overnight at 16 °C. Cells were lysed by sonication in buffer containing 50 mM HEPES pH 7.5, 150 mM NaCl, 2 mM EDTA, 2 mM DTT, 15% (v/v) glycerol with protease inhibitors and centrifuged. GST-Kapβ2 was then purified using Glutathione Sepharose 4B protein purification resin and eluted in buffer described previously with 30 mM glutathione, adjusted to pH 7.5. GST was cleaved using TEV protease overnight at 4 °C or kept on the protein and Kapβ2 was further purified by ion-exchange and size exclusion chromatography in buffer containing 20 mM HEPES pH 7.5, 110 mM potassium acetate, 2 mM magnesium acetate, 2 mM DTT and 10% (v/v) glycerol. Purified proteins were flash-frozen and stored at −80 °C. GST-tagged hnRNPA2 PY-NLS WT and N323Tfs peptides were purified similarly except no ion-exchange step was included.

GST-tagged hnRNPA2 proteins were purified as described^6^. Briefly, WT and mutant hnRNPA2 with N-terminal GST tag were over-expressed into *E. coli* BL21(DE3)-RIL (Invitrogen). Bacteria were grown at 37 °C until reaching an OD600 of ∼0.6 and expression was induced by addition of 1 mM isopropyl 1-thio-β-D-galactopyranoside (IPTG) for 15-18 h at 15 °C. Protein was purified over a glutathione-Sepharose column (GE) according to manufacturer’s instructions. GST-hnRNPA2 was eluted from the glutathione Sepharose with 40 mM HEPES-NaOH, pH 7.4, 150 mM potassium chloride, 5% glycerol, and 20 mM reduced glutathione. Eluted proteins were centrifuged at 16,100 g for 10 min at 4 °C to remove any aggregated material before being flash-frozen in liquid N_2_ and stored at −80 °C. Before each experiment, protein was centrifuged at 16,100 g for 10 min at 4 °C to remove any aggregated material. After centrifugation, the protein concentration in the supernatant was determined by Bradford assay (Bio-Rad) and these proteins were used for aggregation reactions. TEV protease was purified as described ^49^.

### Fluorescence polarization

Synthesized TAMRA-tagged hnRNPA2 PY-NLS WT, N323Tfs, G328Afs, or G331Efs peptide were incubated with increasing amounts of purified Kapβ2 WT in buffer containing 20 mM HEPES pH 7.5, 110 mM potassium acetate, 2 mM magnesium acetate, 2 mM DTT and 10% (v/v) glycerol in Corning 96-well solid black polystyrene plates.

Fluorescence polarization was measured with a Cytation 5 multi-mode plate reader (Biotek) using Gen5 software with 561-nm polarization cube. Analysis was performed using MATLAB. Briefly, fluorescence polarization was converted to anisotropy and K_d_ values were calculated by fitting the resulting curve to the equation 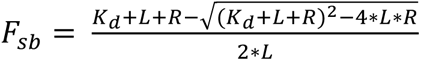; where *F_sb_* is the fraction bound, *L* is the concentration of peptide, and *R* is the concentration of Kapβ2. Each peptide was run in triplicate.

### Generation of *Drosophila* lines and *Drosophila* stocks

The hnRNPA2 WT and D290V *Drosophila* stocks have been previously^6^. Flies carrying pUASTattB-hnRNPA2 N323Tfs, G328Afs, or G331Efs transgenes were generated by a standard injection and φC31 integrase-mediated transgenesis technique (BestGene Inc). GMR-GAL4 was used to express transgenes in eyes; MHC-GAL4 was used to express transgenes in muscle. All *Drosophila* stocks were maintained in a 25 °C incubator with a 12-h day/night cycle.

### Adult *Drosophila* muscle preparation and immunohistochemistry

Adult flies were embedded in a drop of OCT compound (Sakura Finetek) on a slide glass, frozen with liquid nitrogen and bisected sagitally by a razor blade. After fixing with 4% paraformaldehyde in PBS, hemithoraces were stained by Texas Red-X phalloidin (Invitrogen) and DAPI according to manufacturer’s instructions. Stained hemi-thoraces were mounted in 80% glycerol and the musculature was examined by DMIRE2 (Leica, 103). For hnRNPA2 staining, hemithoraces were permeabilized with PBS containing 0.2% Triton X-100 and stained with anti-hnRNPA2B1 (EF-67) antibody (Santa Cruz Biotechnology) and Alexa-488-conjugated secondary antibody (Invitrogen). Stained muscle fibers were dissected and mounted in Fluormount-G (Southern Biotech) and imaged with a Marianas confocal microscope (Zeiss, 363).

### Solubility and biochemical analyses of adult *Drosophila* muscles

Sequential extractions were performed to examine the solubility profile of hnRNPA2. Adult fly thoraces were lysed in cold RIPA buffer (50 mM Tris, pH 7.5, 150 mM NaCl, 1% Triton X-100, 0.5% sodium deoxycholate, 0.1% SDS and 1 mM EDTA) and sonicated. Cell lysates were cleared by centrifugation at 100,000*g* for 30 min at 4 °C to generate RIPA-soluble samples. To prevent carry-overs, the resulting pellets were washed twice with PBS and RIPA-insoluble pellets were then extracted with urea buffer (7 M urea, 2 M thiourea, 4% CHAPS (3-[(3-cholamidopropyl)-dimethylammonio]-1-propanesulphonate), 30 mM Tris, pH 8.5), sonicated and centrifuged at 100,000*g* for 30 min at 22 °C. Protease inhibitors were added to all buffers before use. Protein concentration was determined by the bicinchoninic acid method (Pierce) and proteins were resolved by NuPAGE Novex 4–12% Bis-Tris gels (Invitrogen). For Western blot analysis, thoraces of adult flies were prepared and ground in NuPAGE LDS sample buffer (NP0007, Invitrogen). Samples were then boiled for 5 min and analyzed by standard Western blotting methods provided by Odyssey system (LI-COR) with 4–12% NuPAGE Bis-Tris gels (Invitrogen).

### In vitro fibrillization assays

hnRNPA2 (5 µM) fibrillization (100 µl reaction) was initiated by addition of 1 μl of 2 mg/ml TEV protease in A2 assembly buffer (40 mM HEPES-NaOH, pH 7.4, 150 mM KCl, 5% glycerol, 1 mM DTT, and 20 mM glutathione). The hnRNPA2 fibrillization reactions were incubated at 25 °C for 0-24 h with agitation at 1,200 rpm in an Eppendorf Thermomixer. For sedimentation analysis, at indicated time points, fibrillization reactions were centrifuged at 16,100 g for 10 min at 4 °C. Supernatant and pellet fractions were then resolved by SDS-PAGE and stained with Coomassie Brilliant Blue, and the relative amount in each fraction was determined by densitometry in ImageJ (NIH). All fibrillization assays were performed in triplicate. For electron microscopy, fibrillization reactions (10 μl) were adsorbed onto glow-discharged 300-mesh Formvar/carbon coated copper grids (Electron Microscopy Sciences) and stained with 2% (w/v) aqueous uranyl acetate. Excess liquid was removed, and grids were allowed to air dry. Samples were viewed on a JEOL 1010 transmission electron microscope.

---

**Supplementary table 1:**
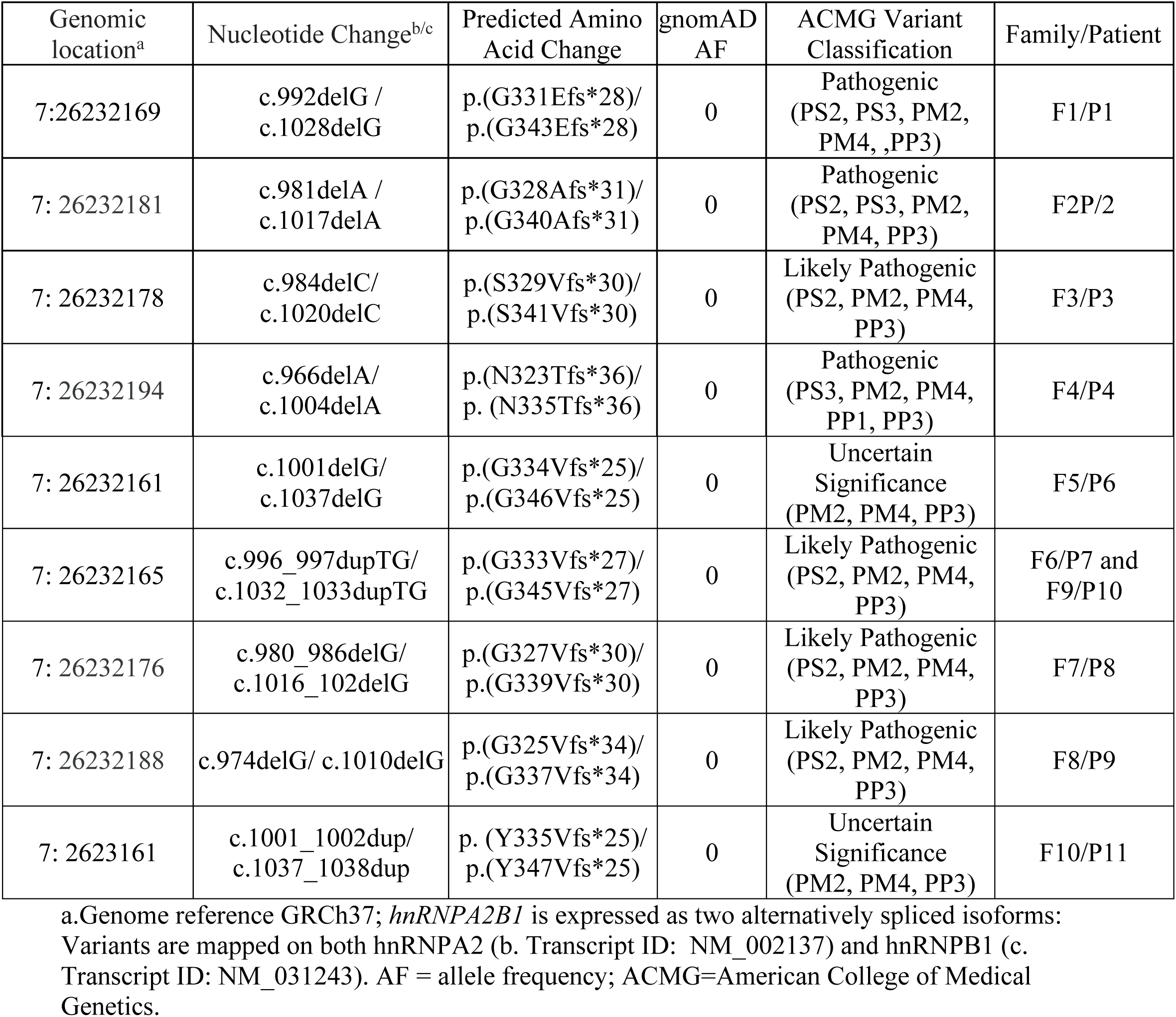
*hnRNPA2B1* variants identified in the cohort.

| Genomic location<sup>a</sup> | Nucleotide Change<sup>b/c</sup> | Predicted Amino Acid Change | gnomAD AF | ACMG Variant Classification | Family/Patient |
| --- | --- | --- | --- | --- | --- |
| 7:26232169 | c.992delG / c.1028delG | p.(G331Efs*28)/ p.(G343Efs*28) | 0 | Pathogenic (PS2, PS3, PM2, PM4, ,PP3) | F1/P1 |
| 7: 26232181 | c.981delA / c.1017delA | p.(G328Afs*31)/ p.(G340Afs*31) | 0 | Pathogenic (PS2, PS3, PM2, PM4, PP3) | F2P/2 |
| 7: 26232178 | c.984delC/ c.1020delC | p.(S329Vfs*30)/ p.(S341Vfs*30) | 0 | Likely Pathogenic (PS2, PM2, PM4, PP3) | F3/P3 |
| 7: 26232194 | c.966delA/ c.1004delA | p.(N323Tfs*36)/ p. (N335Tfs*36) | 0 | Pathogenic (PS3, PM2, PM4, PP1, PP3) | F4/P4 |
| 7: 26232161 | c.1001delG/ c.1037delG | p.(G334Vfs*25)/ p.(G346Vfs*25) | 0 | Uncertain Significance (PM2, PM4, PP3) | F5/P6 |
| 7: 26232165 | c.996_997dupTG/ c.1032_1033dupTG | p.(G333Vfs*27)/ p.(G345Vfs*27) | 0 | Likely Pathogenic (PS2, PM2, PM4, PP3) | F6/P7 and F9/P10 |
| 7: 26232176 | c.980_986delG/ c.1016_102delG | p.(G327Vfs*30)/ p.(G339Vfs*30) | 0 | Likely Pathogenic (PS2, PM2, PM4, PP3) | F7/P8 |
| 7: 26232188 | c.974delG/ c.1010delG | p.(G325Vfs*34)/ p.(G337Vfs*34) | 0 | Likely Pathogenic (PS2, PM2, PM4, PP3) | F8/P9 |
| 7: 2623161 | c.1001_1002dup/ c.1037_1038dup | p. (Y335Vfs*25)/ p.(Y347Vfs*25) | 0 | Uncertain Significance (PM2, PM4, PP3) | F10/P11 |

a.Genome reference GRCh37; *hnRNPA2B1* is expressed as two alternatively spliced isoforms: Variants are mapped on both *hnRNPA2* (b. Transcript ID: NM\_002137) and *hnRNPB1* (c. Transcript ID: NM\_031243). AF = allele frequency; ACMG=American College of Medical Genetics.

